# SEIR-Based Modelling of COVID-19 Spread in Malaysia: A Mathematical Approach to Public Health Interventions and Optimal Control Solutions

**DOI:** 10.1101/2025.06.27.25330398

**Authors:** Hakimah Yusop, Sharifa Ezat Wan Puteh, Mohd Almie Alias

## Abstract

The COVID-19 pandemic has posed unprecedented challenges to public health systems globally, requiring the adoption of various control measures to mitigate viral spread. This study aims to model the transmission dynamics of COVID-19 in Malaysia and evaluate the effectiveness of key intervention strategies using a SEIR-based mathematical model, augmented by optimal control theory. The model incorporates epidemiological data from the second wave of COVID-19 in Malaysia, considering critical parameters such as the transmission rate (β = 8.00373) and latency period (α = 0.175), including the key public health interventions, derived through curve fitting to the observed data. The model revealed that the nationwide Movement Control Orders (MCO) had led to a substantial reduction in the contact rate (ƈ) from 1.0 to about 0.3, self-protection measures including the use of face masks further reduced transmissibility by 0.01507, with screening and contact tracing rates exceeding 90% while case detection rate was about 0.3.

To optimize intervention strategies, optimal control theory was employed, aiming to balance the effectiveness of controlling the virus with minimizing the socio-economic costs of these interventions. Several simulations were conducted to explore the impact of varying control measures, including enhanced screening and contact tracing, case detection, movement control, and self-protection. Simulations revealed that movement control was most effective in the early stages but should not be prolonged to minimize economic disruptions. Meanwhile, simulations also underscored the importance of self-protection, enhanced screening and contact tracing, and case detection, highlighting their critical role in long-term pandemic control. Results from the simulations also emphasized the importance of flexible, adaptive intervention strategies that can be tailored to the evolving situation.

This study highlights the utility of mathematical modelling in understanding COVID-19 transmission and guiding public health responses. The findings from this study provide valuable insights into the optimal allocation of resources and the design of adaptive strategies to control COVID-19 and future pandemics. The use of optimal control modeling in this context highlights its utility for guiding evidence-based decision-making, offering a framework for improving pandemic preparedness and response.

## 1.0 Introduction

The emergence of COVID-19, a severe and highly transmissible respiratory illness, has precipitated one of the most profound global public health crises in modern history. First identified in Wuhan, China, in late 2019, the novel coronavirus (SARS-CoV-2) rapidly spread across continents, overwhelming healthcare systems, disrupting global economies, and inflicting significant social, psychological, and political repercussions ^12^. As the pandemic unfolded, it exposed critical vulnerabilities in both healthcare infrastructure and pandemic preparedness across the world. Despite a series of interventions and containment efforts as recommended by the World Health Organization (WHO), the virus proved to be difficult to control, leading to repeated waves of outbreaks, underscoring the complexity of pandemic management^3^.

Malaysia, like many other nations, confronted the dual challenge of curbing the spread of the virus while simultaneously managing the socio-economic repercussions of prolonged public health restrictions. Early in the pandemic, in the absence of vaccines or specific treatments, the Malaysian government responded with a combination of stringent measures such as widespread lockdowns, travel restrictions, mass testing, and contact tracing ^4,5^. While these efforts achieved some success in reducing transmission, the situation required constant adaptation as infection rates fluctuated and more transmissible variants emerged ^6–8^. The strain on the public health system and the broader economic impacts—including disruptions to industries, widespread job losses, and declines in productivity—were profound, exacerbating the long-term effects on Malaysia’s financial stability ^9,10^.

Given the highly unpredictable nature of emerging infectious diseases, such as COVID-19, and the rapidly evolving global landscape of pandemics, there is an urgent need for effective decision-making tools in public health ^11,12^. Mathematical modeling has emerged as an invaluable tool in this context, enabling the simulation of disease dynamics and the evaluation of various intervention strategies ^13^. In particular, compartmental models like the SEIR (Susceptible, Exposed, Infectious, Recovered) model have been widely used to estimate key epidemiological parameters, predict infection trends, and assess the potential impact of public health measures ^14–16^. These models provide an evidence-based approach to understanding the complexities of disease transmission and have proven instrumental in informing public health policy during pandemics.

This study develops a SEIR-based epidemiological model to examine the transmission dynamics of COVID-19 in Malaysia, with particular emphasis on assessing the effectiveness of early intervention strategies. The absence of vaccines or targeted treatments during the initial stages of the pandemic presented a significant challenge to public health authorities, necessitating the implementation of non-pharmaceutical interventions to control viral spread. By simulating various scenarios and incorporating key parameters, this model aims to provide a comprehensive understanding of COVID-19’s characteristics in Malaysia. Moreover, the study seeks to evaluate how public health interventions—such as movement restrictions, testing, and quarantine measures—shaped the trajectory of the outbreak.

A central feature of this study is the application of optimal control theory, a mathematical framework used to determine the most effective strategies for managing the pandemic while minimizing the associated costs. By formulating the problem as an optimal control problem, the study seeks to identify the time-dependent controls— such as screening and contact tracing, social distancing, and self-protection measures—that can most effectively reduce the disease burden. This approach allows policymakers to evaluate the trade-offs between public health benefits and the socio-economic costs of intervention measures.

Through the integration of optimal control theory with epidemiological modeling, this study aims to provide critical insights into how early and adaptive public health measures can mitigate the spread of COVID-19, preserve healthcare resources, and minimize the societal impact of pandemics. The findings from this research will offer a framework for public health officials and policymakers to optimize their response strategies, not only in the context of COVID-19 but also for future infectious disease outbreaks. By highlighting the relationships between intervention measures and disease transmission dynamics, the model serves as a valuable tool for informed decision-making in pandemic preparedness and response.

## 2.0 Methods

### 2.1 Model Formulation

To capture the unique dynamics of COVID-19 transmission in Malaysia, an extended SEIR model was developed. This model accounts for the epidemiological conditions and public health interventions specific to Malaysia during the pandemic. The total population, denoted by (*N*), is divided into several compartments: susceptible (S), exposed (E), infectious (I), recovered (R), and death due to COVID-19 (D). Each exposed (E) and infectious (I) compartments are further subdivided to differentiate between quarantined (E_Q_ and I_Q_) and non-quarantined (E_X_ and I_X_) individuals, accounting for public health interventions which are quarantine and case isolation.

As in the basic SEIR model, the susceptible (S) compartment consists of all susceptible individuals who are at risk of being exposed to the infection. Individuals who have been exposed to the infection due to contact with an infected person, but the virus is still latent and thus cannot infect others, are categorized in the exposed (E) compartment. Movement of individuals from the susceptible (S) to the exposed (E) compartment is driven by the force of infection, which depends on the disease transmission rate β, and the effective daily contact rate between infectious (I) and susceptible (S) individuals. Physical distancing and movement restrictions can reduce disease transmission by decreasing the daily contact rate, represented by the parameter ƈ. The daily contact rate was assigned to a value between 0 and 1, where 0 indicates no contact at all, and 1 signifies maximum contact, reflecting normal social interactions. The model also incorporates self-protection behaviors, such as good hygiene, regular hand washing, use of hand sanitizers, and disinfection practices, which modify the force of infection through parameter ᴋ, ranging from 0 (no self-protection) to 1 (complete self-protection).

Enhanced screening and contact tracing, represented by the parameter τ, enable the identification and quarantine of at-risk individuals before their transition into compartment I, thereby reducing infection risk. Quarantined individuals are categorized into E_Q_, while those who remain undetected (1-τ) are classified into E_X_.

Following the latency period, individuals in E transition to I (Infectious). This transition is represented by the parameter α, which is the reciprocal of the latent period. Quarantined individuals in compartment E_Q_ are assumed to remain isolated until the end of the latency period, after which they move directly into compartment I_Q_. Meanwhile, individuals in compartment E_X_ transition into compartment I_X_ after completing the latency phase. Individuals in compartment I are those who have become infected and are capable of transmitting the disease. However, due to isolation of COVID-19 positive patients, it is assumed that only undetected infectious individuals (I_X_) spread the disease in the community.

The I_D_ compartment is added to the model structure to represent individuals diagnosed as confirmed COVID-19 positive cases, allowing the real-world data on COVID-19 cases in Malaysia to be incorporated into the mathematical system of equations. Parameter Δ represents the case detection rate, where Δ_Q_ represents the case detection rate among quarantined individuals, while Δ_X_ denotes the detection rate among individuals in I_X_.

Infected individuals will either recover and gain immunity, classified into compartment R (Recovered), or succumb to COVID-19, categorized in compartment D (Death). The recovery rate is represented by parameter γ, while the mortality rate is represented by the parameter μ. To account for variations in recovery rates between untreated individuals and those diagnosed and receiving treatment or healthcare, parameters γ_D_ and μ_D_ are used to represent the recovery and mortality rates among diagnosed individuals. Recovered individuals may become reinfected due to waning of immunity over time, represented by the parameter ω_R_.

The transmission dynamics of COVID-19 within the Malaysian population, as described, are illustrated in Figure 1. The model’s structure is designed to provide insight into the effects of interventions measures on the spread of COVID-19 in Malaysia, offering a framework for public health decision-making and resource allocation.

**Figure 1.**
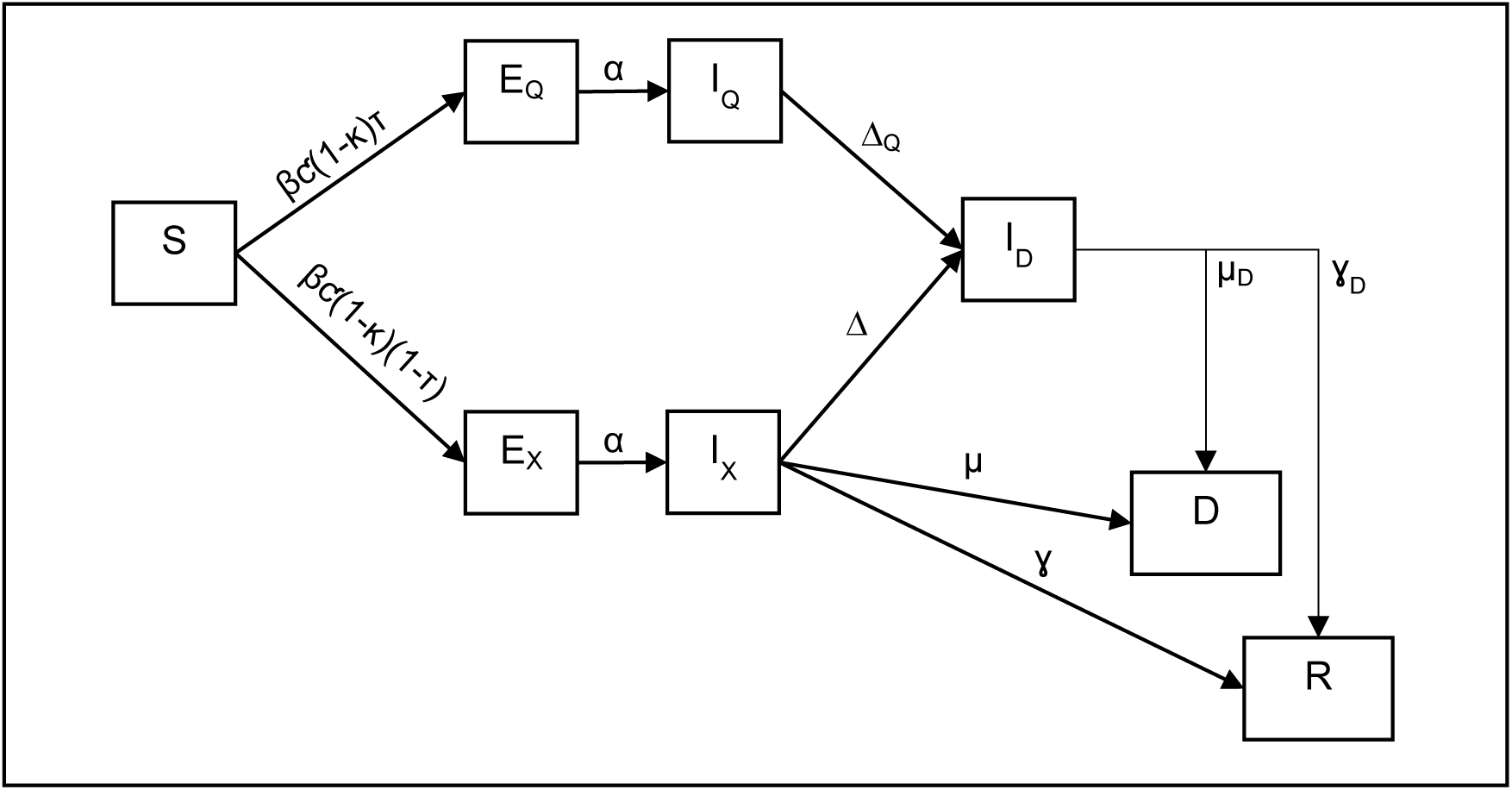
Illustration of SEIR-Based Model Structure for the COVID-19 Transmission Dynamics in Malaysia During The Initial Phase of Pandemic

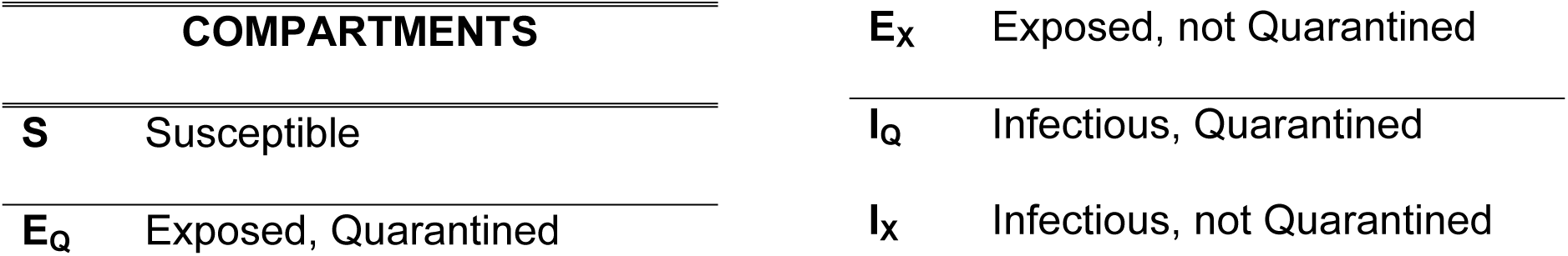
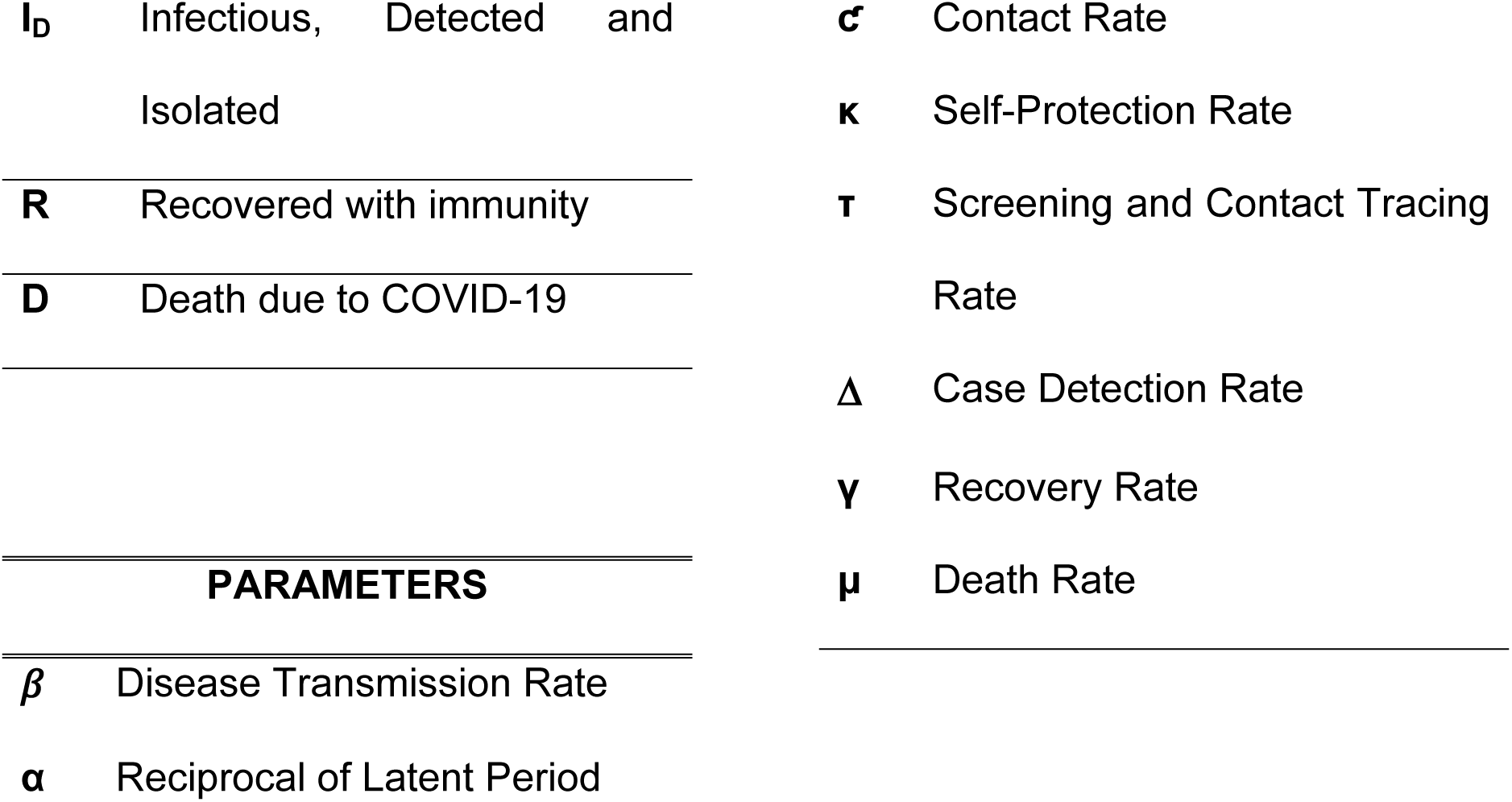

The corresponding system of differential equations underlying the extended SEIR model are as follows:

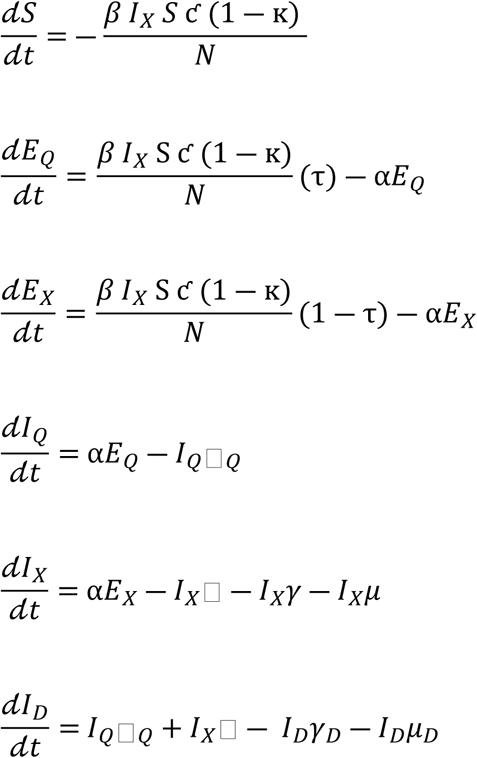

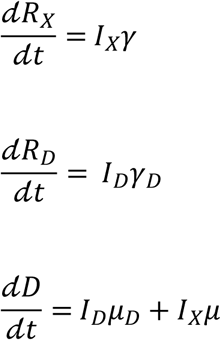

### 2.2 Modelling Tool and Solution

The model development, numerical solution, and data analysis were conducted using Python, leveraging its rich ecosystem of scientific libraries. The system of differential equations of the model was solved through mathematical numerical integration, utilizing the odeint() function from the SciPy library. To ensure the model closely represented real-world dynamics, parameter estimation was conducted using the least squares method achieved using the curve_fit() function, also from the SciPy library, which iteratively adjusted the model parameters to best match the real COVID-19 transmission data.

The data used for model calibration and analysis were sourced from the official open-data repository of the Ministry of Health Malaysia (MoH), available on their GitHub account accessible via https://github.com/MoH-Malaysia/covid19-public. This repository provided daily updates on key epidemiological indicators, including the number of active cases, recoveries, and deaths. The model was calibrated specifically to represent Malaysia’s second wave of COVID-19, spanning from February 27, 2020, to September 1, 2020.

The curve fitting process was conducted by aligning the active case data with the I_D_ (infectious detected) compartment, recovered case data with the R_D_ (recovered) compartment, and COVID-19 mortality data with the D (death) compartment, for the entire second wave of COVID-19 in Malaysia to estimate the parameters of disease characteristics, which are the transmission rate (β), reciprocal of the latent period (α), and recovery rate (ɣ). The first wave was excluded from the analysis as it primarily involved sporadic cases, without sustained community transmission. Additionally, curve fitting was performed stratified according to the different phases of the Malaysian government’s intervention measures to account for the impact of these control measures on the transmission dynamics. The critical intervention milestones considered in this analysis are as follows:

- February 27, 2020: Onset of the second wave of COVID-19 in Malaysia.
- March 18, 2020: First day of the implementation of Movement Control Order (MCO), followed by Enhanced Movement Control Order (EMCO).
- May 4, 2020: First day of the implementation of Conditional Movement Control Order (CMCO).
- June 10, 2020: End of the Movement Control Order (MCO).
- August 1, 2020: Introduction of mandatory mask-wearing in public spaces.

These intervention phases represent the core components of Malaysia’s public health strategy in mitigating the spread of COVID-19. The phase-specific curve-fitting approach enabled a detailed assessment of how each intervention influenced disease transmission and progression. By comparing parameter estimates across these phases, the model provided insights into the effectiveness of the public health measures, elucidating how policy-driven behavioral changes contributed to controlling the pandemic’s trajectory within the country.

### 2.3 Optimal Control Formulation

Optimal control modelling approach is adopted to determine the most effective strategy to implement until vaccine deployment.

In formulating the optimal control model system, the extended SEIR-based model is restructured into an optimal control model with four time-dependent controls as follows:

(i) u_τ_(t) - control variable for enhanced screening and contact-tracing followed by quarantine at risk individuals
(ii) u_Δ_(t) - control variable for case detection followed by isolation of the diagnosed COVID-19 infected cases
(iii) u_ƈ_(t) - control variable for social distancing measures and movement control orders
(iv) u_ᴋ_(t) - control variable for self-protection measures aimed at decreasing the transmission rate

These controls are given value ranging from 0 indicating no control, to 1 indicating full control.

The resulting control model is formulated via the following system:

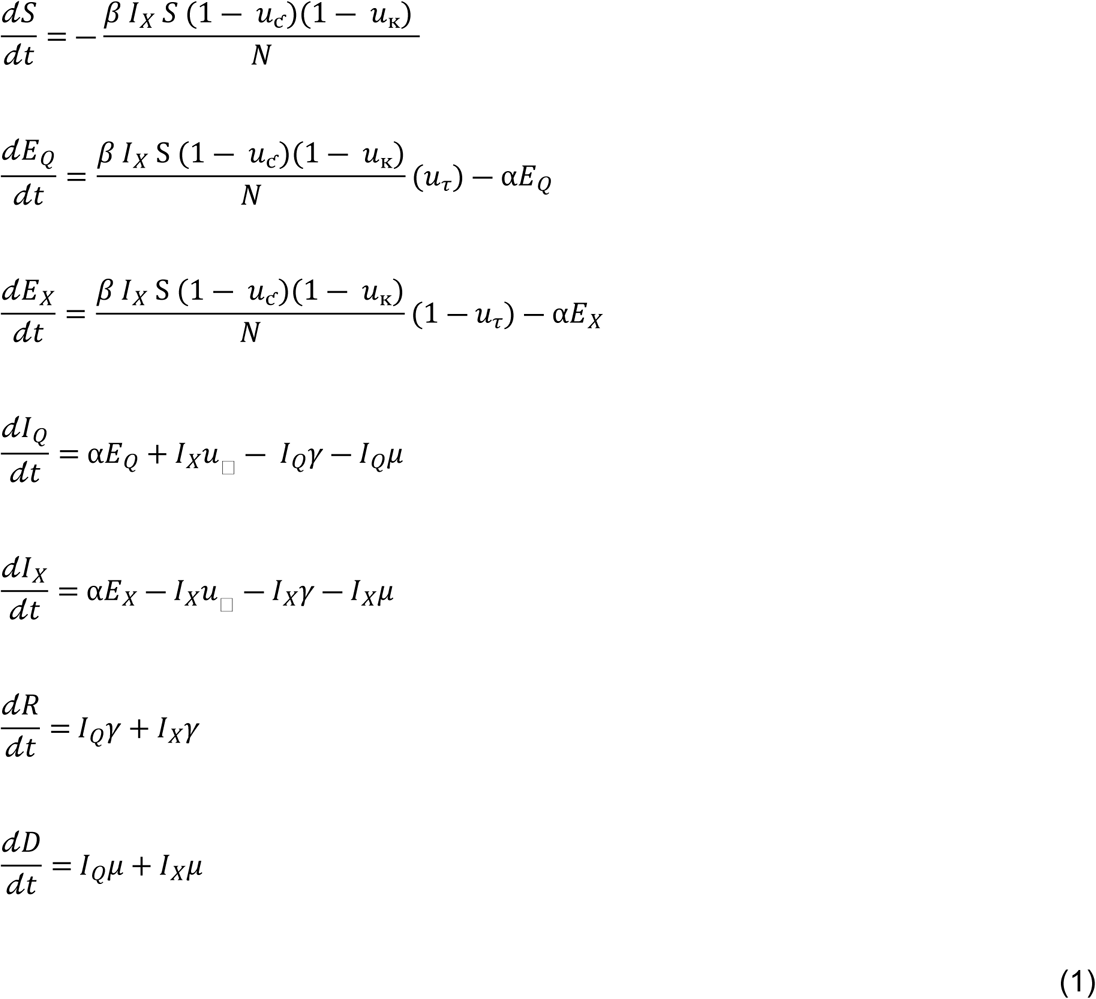

The objective of this optimal control problem is to identify the optimal control strategy over time that minimises the burden of disease. The burden of disease is defined as the cumulative sum of the costs associated with the total number of infectious individuals and the cumulative costs of interventions. Therefore, the functional objective is written as:

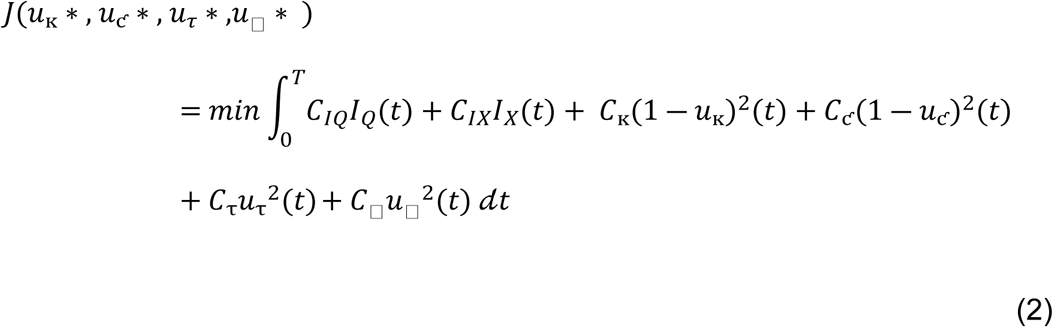

The constants C_IQ_ and C_IX_ are weights related to the infectious, and undetected infectious individuals, respectively. While, the weights C_ᴋ_, C_ƈ_, C_τ_ and C_Δ_ are in association with the time-dependent control functions u_ᴋ_, u_ƈ_, u_τ_ and u_Δ_, respectively.

### 2.4 Optimal Control Characterization

Pontryagin’s Maximum Principle ^17^ is used to form the necessary conditions that an optimal control and state must satisfy. The Hamiltonian for the above optimal control system is defined by:

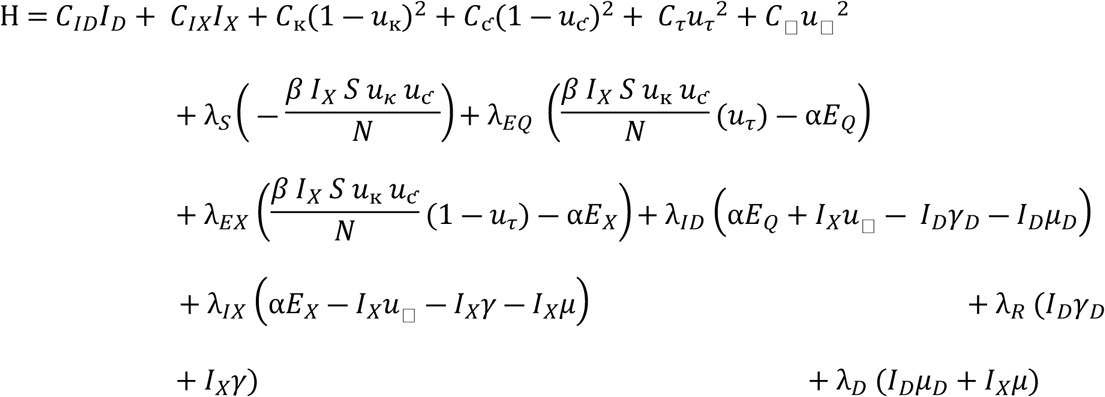

The adjoint system satisfying the condition (2) is as below:

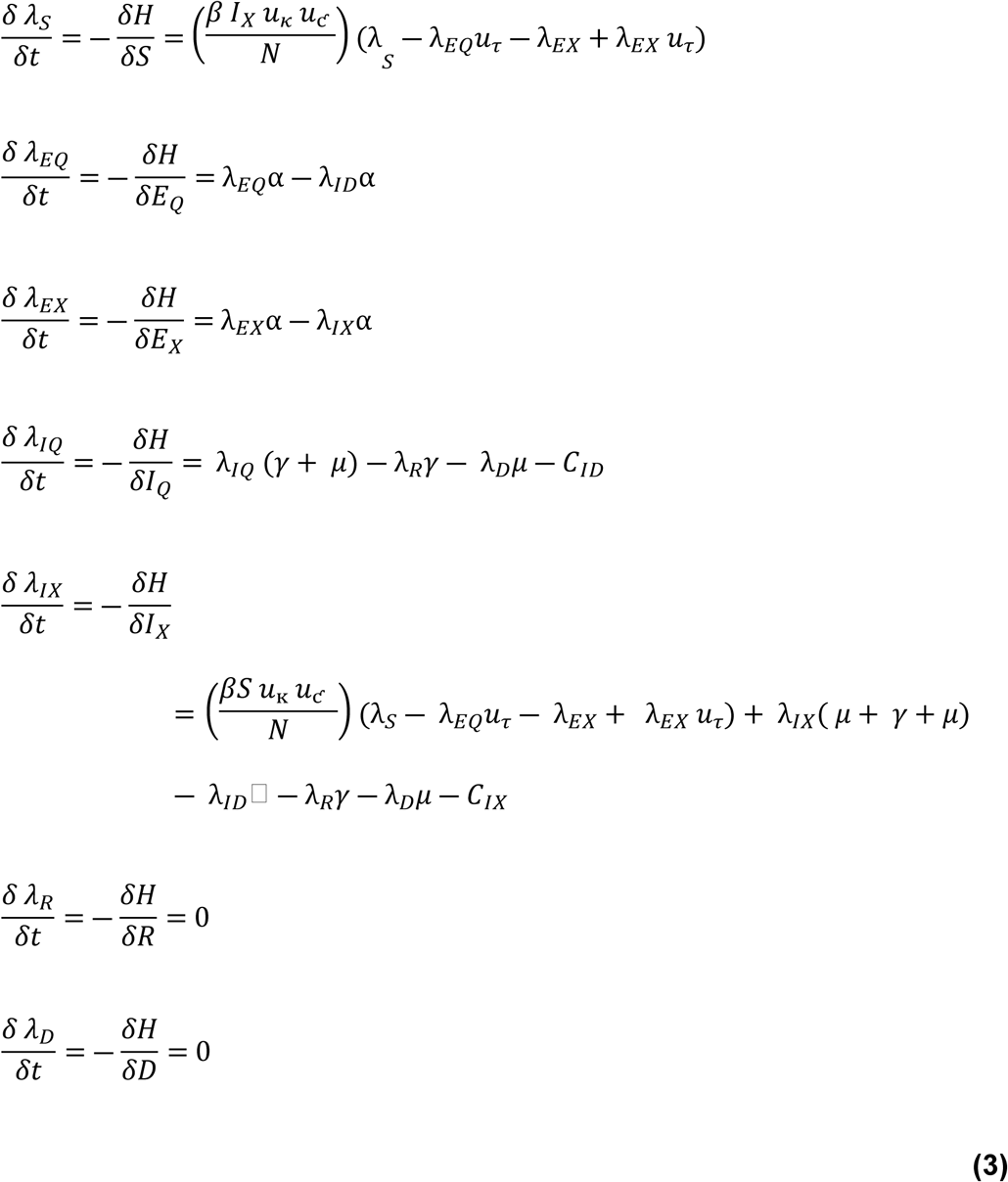

Control functions (*u*_ᴋ_ ∗, *u*_ƈ_ ∗ , *u*_*τ*_ ∗ ,*u*_□_ ∗) which satisfy the optimality condition are given by:

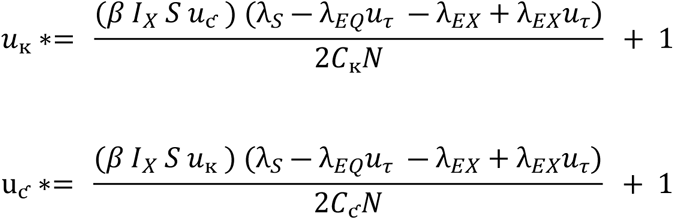

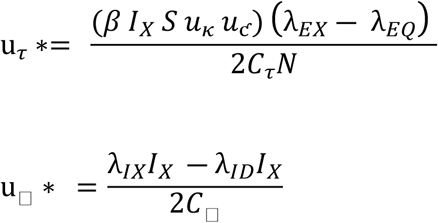

Parameter values obtained from model fit are applied as the bounds on the controls, as following:

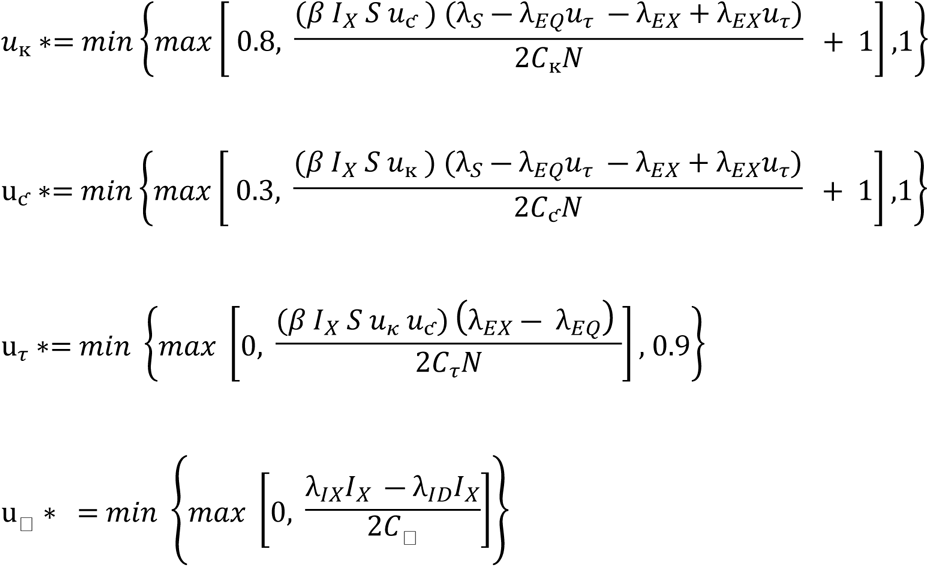

### 2.4 Optimal Control Solutions

The optimal control problem, including the adjoint equation and the optimal control characterization are solved numerically using the Runge-Kutta order four scheme (RK4). The results are presented graphically. Parameter values obtained from the previous curve fitting, as listed in Table 1, are used in the simulation process. The results are analysed in the context of COVID-19 pandemic in Malaysia.

**Table 1.**
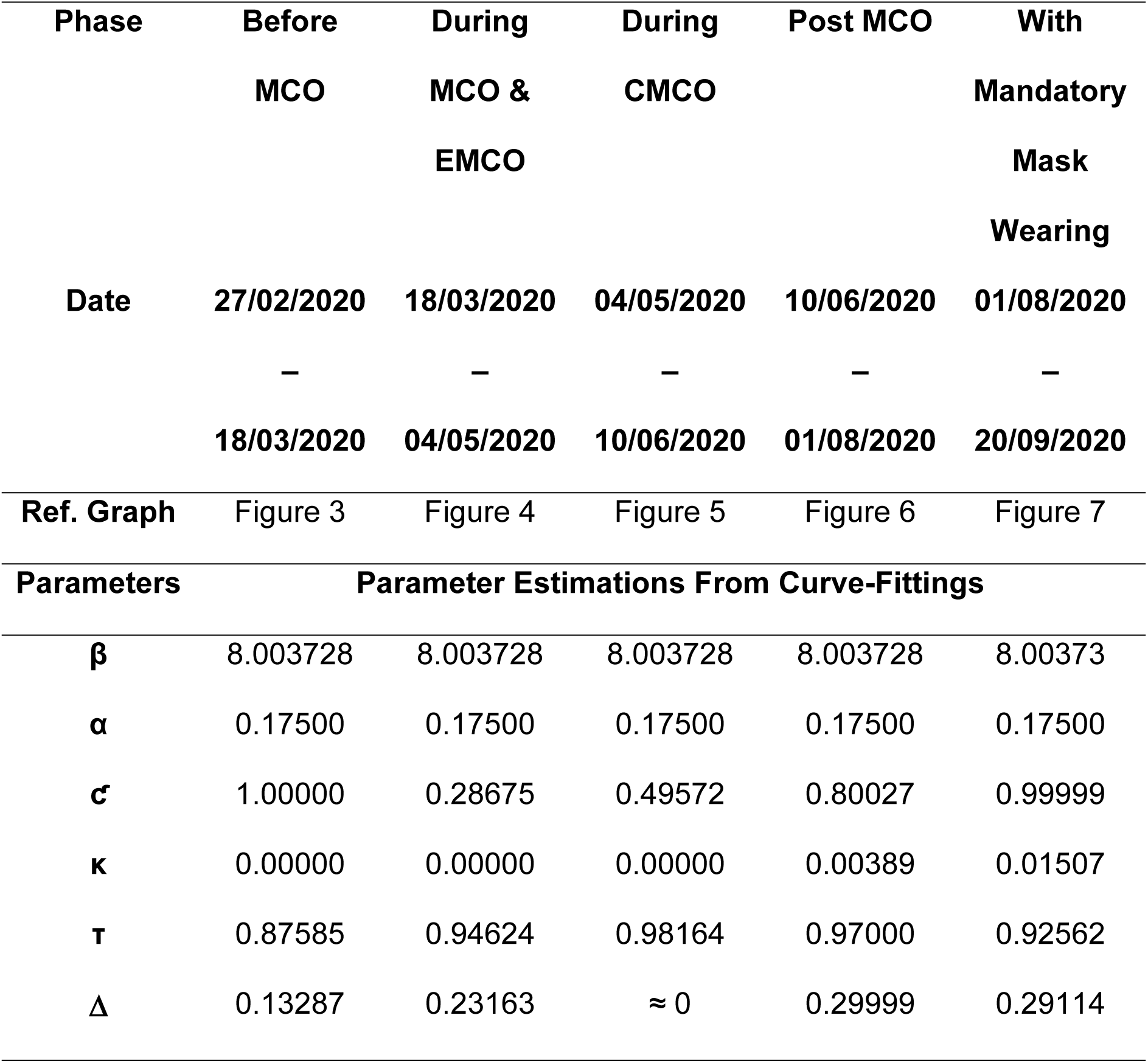
Table of Parameter Estimation Values Obtained From Curve-Fitting.

## 3.0 Results and Discussion

### 3.1 Model Calibration and Parameter Estimations For The Characteristics of Disease

The calibration of the model to the second wave of COVID-19 in Malaysia reveals key insights into the disease’s dynamics during the early phases of the pandemic. **Error! Reference source not found.** illustrates the curve-fitting process, where the model’s best fit to the real-world data is shown. From this process, several important parameters were estimated. The transmission rate (β) was 8.00373, indicating a rapid spread of the virus in the early phase of the second wave. The reciprocal of the latency period was found to be 0.175 per day, translating to a latent period of approximately 5.7 days. This result closely aligns with a retrospective study conducted across eight regions in China from July 10, 2020, to April 2, 2021, which found an overall average latency period of 5.5 days. The latency period for symptomatic cases was similarly 5.5 days, while for asymptomatic cases was 5.2 days. The average latency period observed in that study was 1.4 days shorter than the incubation period, which was estimated to be 6.9 days ^23^. This latency period is also consistent with a study conducted in Anhui Province, China, where the median incubation period was 5.5 days ^18^. The transmission rate and latency period were assumed to remain constant throughout this wave, as no new variants emerged that could potentially alter the disease’s characteristics. The average recovery rate, denoted by γ, was 0.05027 per day, corresponding to an average recovery time of about 20 days. This parameter is crucial in understanding the burden on healthcare systems. took approximately 20 days to recover from the infection. The COVID-19 mortality rate, represented by μ, was 0.00079 per day. These parameter estimates offer insights into the transmission dynamics and clinical outcomes of COVID-19 during this period, reflecting the pre-vaccination phase and before the emergence of significant viral mutations that might affect disease transmissibility or virulence.

In this mathematical modelling, transmission rate (β) is prioritized over the basic reproduction number (R_0_) as a more stable and informative measure for assessing the spread of virus. While the basic reproduction number fluctuates with varying external factors (e.g., population density and behaviour), β provides a clearer assessment of disease spread, particularly when factoring in interventions like physical distancing and self-protection behaviours ^19,20^. These interventions can be directly observed in the model through parameters like movement control order (ƈ), self-protection (ᴋ), and case isolation ^21^. This approach is employed in this study to illustrate the impact of disease control interventions on disease trends.

In epidemiological models, the E compartment accurately represents individuals in the latency phase, with the parameter α signifying the transition from the exposed to the infectious phase. Understanding the latency period is crucial for designing appropriate public health strategies. The relatively short latency period supports the need for rapid response strategies like frequent screening and contact tracing. These strategies are essential for interrupting transmission chains and preventing large-scale outbreaks. The latency period plays a significant role in disease dynamics, making the SEIR model, with its inclusion of the E compartment, highly appropriate and accurate for modelling COVID-19 transmission.

The recovery rate reflects the transition rate of individuals from the Infectious (I) compartment to the Recovered (R) compartment, which can also be defined as the infectious period. This parameter is a crucial indicator of healthcare system capacity during a pandemic. A faster recovery rate implies a more efficient turnover of patients in hospitals, particularly in critical care units, allowing healthcare facilities to accommodate more patients during surges. Conversely, a slower recovery rate may signal an impending strain on healthcare resources, requiring authorities to expand hospital capacity, increase ICU bed availability, and recruit additional healthcare personnel. Understanding the recovery dynamics not only helps manage healthcare resources but also provides a more accurate projection of disease burden, essential for guiding public health responses and strategic planning.

### 3.2 Calibration of Parameters Related to Intervention Strategies

Figure 3 through Figure 7 illustrate the model’s curve-fitting process across different phases of the second wave, corresponding to varying intervention measures implemented by the Malaysian government, which were key factors in reducing the transmission rate, while Table 1 presents the corresponding parameter estimations.

**Figure 2.**
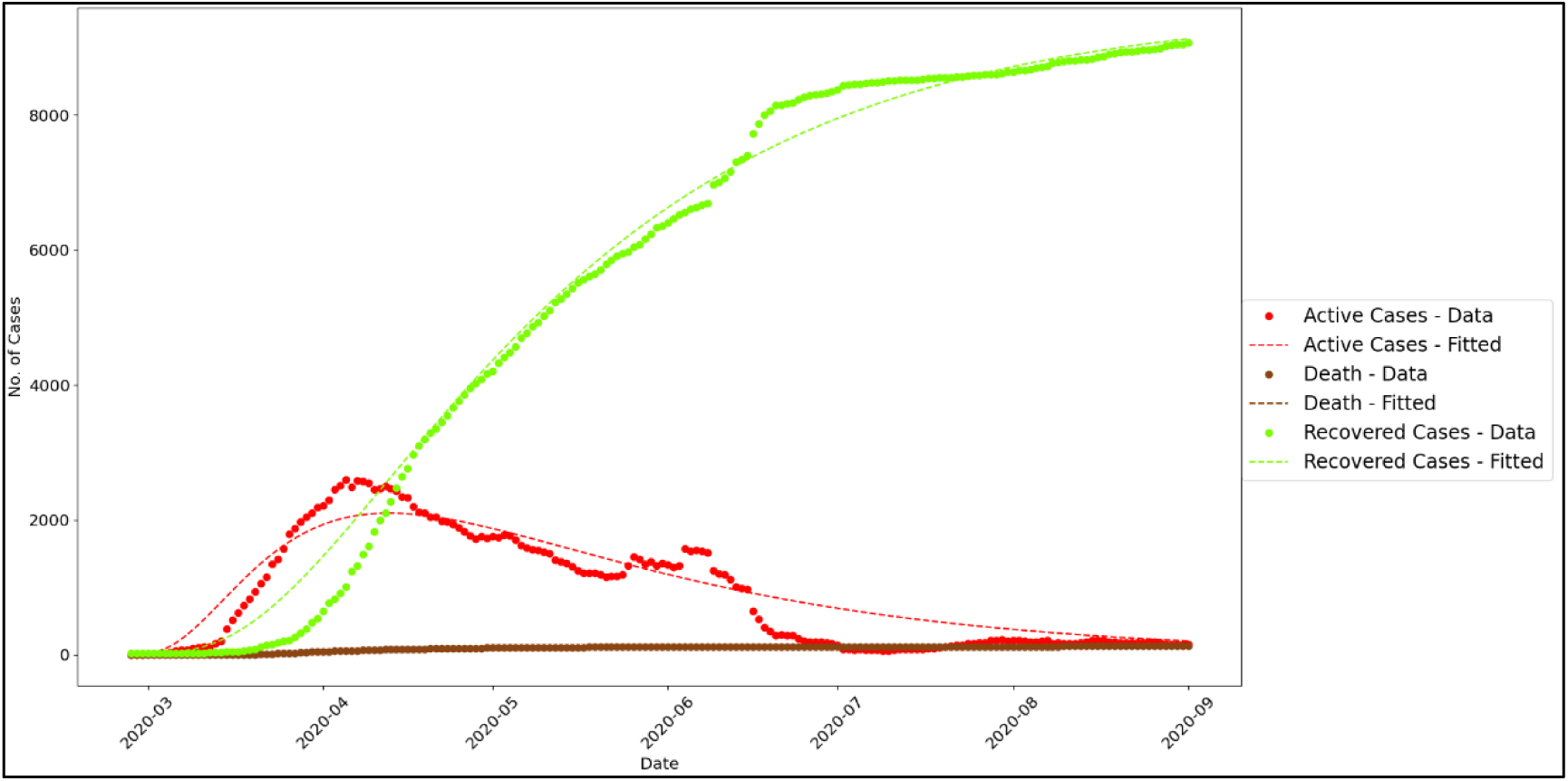
Curve Fitting Graph of The Second Wave of COVID-19 in Malaysia

**Figure 3.**
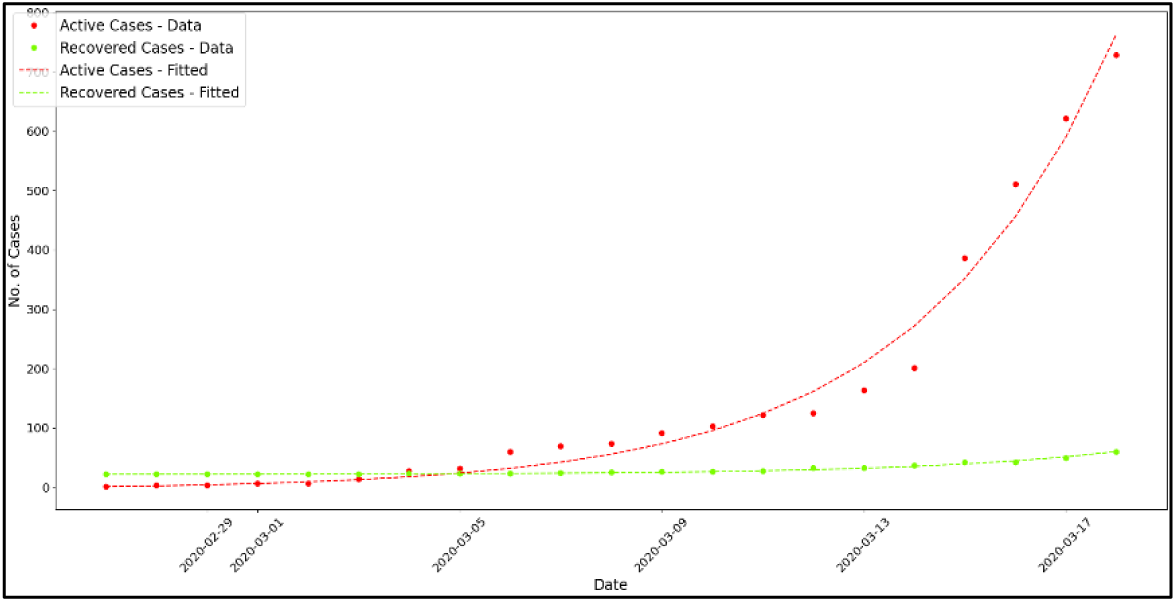
Curve Fitting Graph During The Initial Phase Before MCO

**Figure 4.**
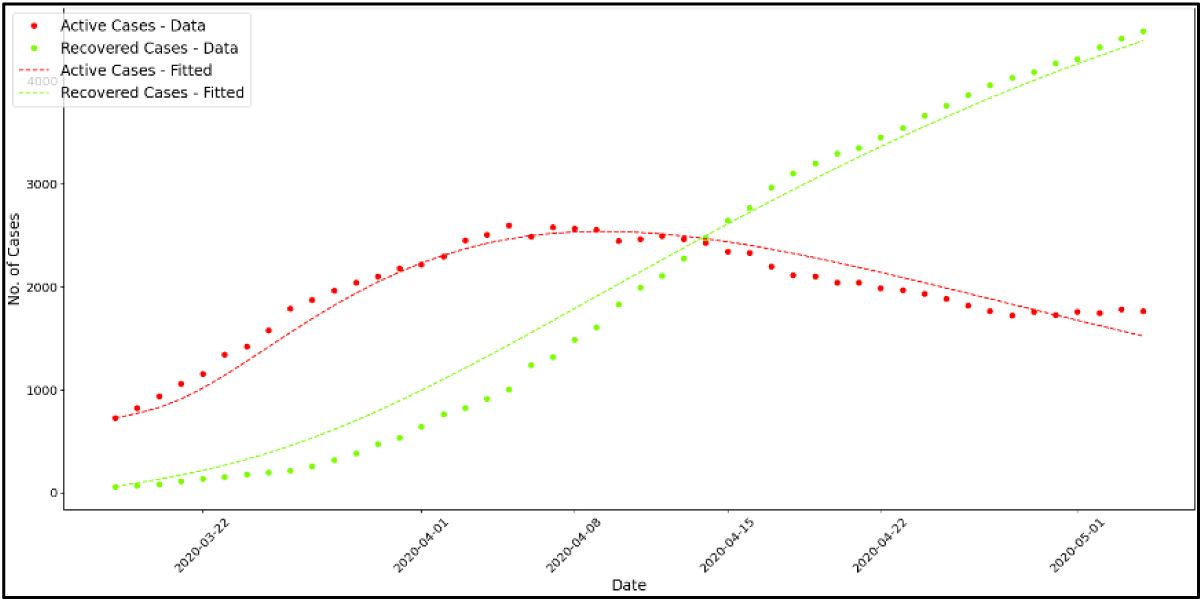
Curve Fitting Graph During MCO & EMCO

**Figure 5.**
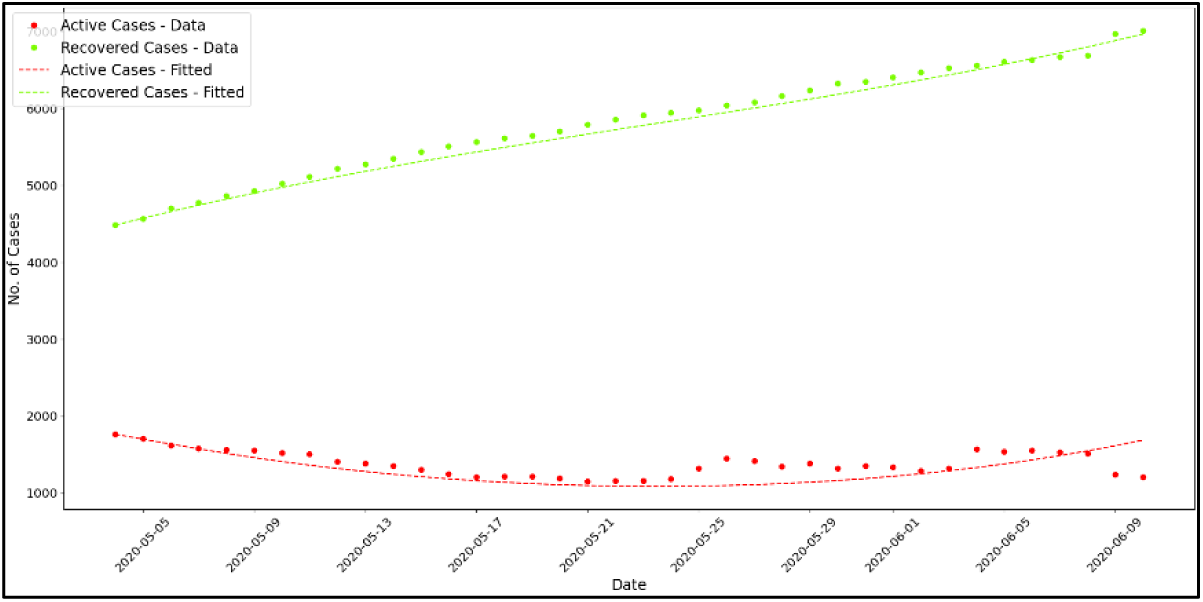
Curve Fitting Graph During CMCO

**Figure 6.**
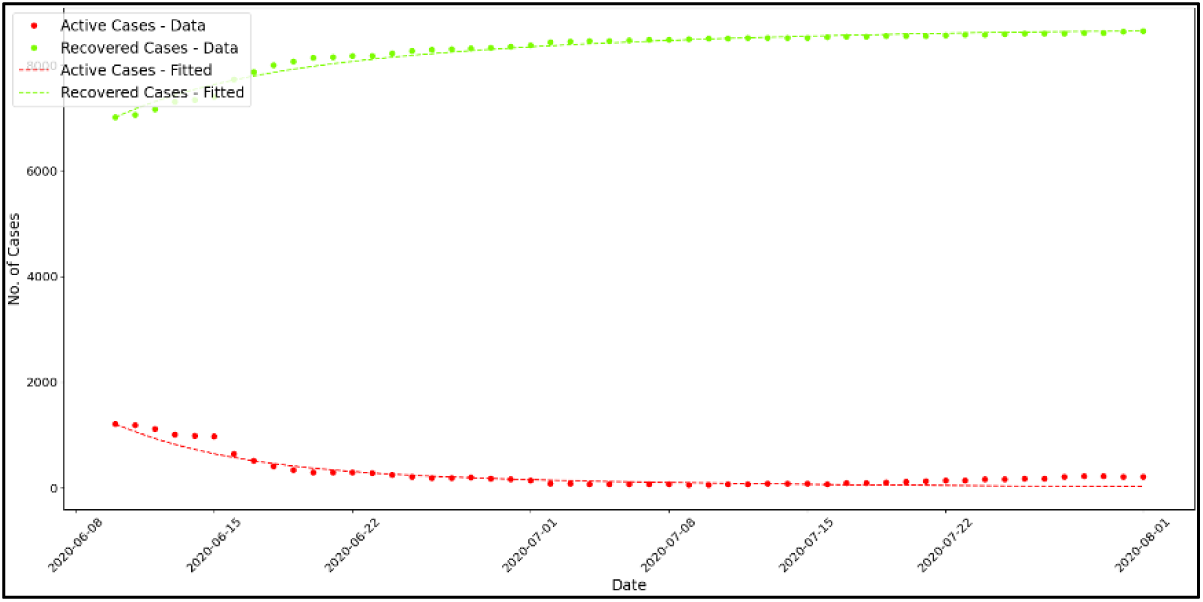
Curve Fitting Graph Without MCO

Through this modelling, the impact of movement control interventions on disease trends is captured using parameter ƈ, which serves as a coefficient for changes in the force of transmission. At the onset of the second wave of COVID-19 in Malaysia, before the implementation of Movement Control Order (MCO), the contact rate ƈ was set to 1.0, representing normal social interactions. The effect of MCO on the force of infection is reflected in the change of parameter ƈ from 1.0 to 0.28675 during the MCO and Enhanced MCO (EMCO) periods, and increased slightly to 0.49572 during Conditional MCO (CMCO). During the phase without movement restrictions, parameter ƈ initially increased to 0.80027 which is still lower compared to the early phase of COVID-19 emergence, and approached 1 (0.99999) during the early phase of third-wave equivalent to the normal contact rate in the Malaysian population.

Initially set to 0.0 indicating no self-protection, the estimated value of parameter ᴋ representing self-protection after movement control measures were lifted and before mandatory use of face masks was 0.00389. This value is assumed to represent self-protection measures such as hygiene practices, frequent handwashing, and the use of hand sanitizers. This parameter value results in a reduction in the force of infection by a coefficient of 1 - 0.00389, which equals to 0.99611. After mandatory use of face masks was implemented, curve fitting indicated that parameter ᴋ increased to 0.01507. This parameter then increased further during the third wave, reaching a peak of 0.27648, indicating widespread adoption of self-protection behaviours.

The rate of enhanced screening and contact tracing followed by quarantine represented by parameter τ was found to exceed 0.9 per day or 90% during the second and third waves, underscoring the importance of early case detection and quarantine measures in controlling the pandemic. Only at the beginning of the second wave did the screening and contact tracing rate decrease slightly; however, it remained above 0.85 per day or 85%. Notably, even during periods of lower contact tracing rates, this intervention remained critical in containing the virus’s spread.

The value of parameter Δ, which represents the case detection rate followed by isolation, was found to range between 0.13 and 0.3 per day during the second wave and early phase of third wave. However, the value of parameter Δ approached zero when the COVID-19 trend plateaued between May and June 2020. Similarly, in the early phase of the third wave, the value of parameter Δ was also found to approach zero. Parameter Δ subsequently was observed to increase, approaching 0.5 per day after November 2020, during the third wave of COVID-19 in Malaysia.

The findings of this study underscore the critical role of disease control interventions—such as quarantine, isolation, movement restrictions, mask mandates, and self-protection measures—in mitigating COVID-19 transmission. By accurately modelling these interventions, public health authorities can better plan and implement targeted strategies to control outbreaks and allocate resources effectively.

### 3.2 Optimal Control

#### Simulation 1: All Four Controls (u_ᴋ_, u_ƈ_, uτ, u_Δ_), with Costs Approximating Real Costs

To obtain the optimal control for a situation close to the real-world scenario, all weights in the objective function were set to values that closely match the actual costs, while the maximum control rates were set as determined from the curve fitting adjustments in the previous section. The weights C_ID_ and C_IX_ were set to 4,000, while the weight C**_D_** was set to 10,000,000, corresponding to the cost of treatment for one infected person at RM4,000 per day and the cost of each COVID-19-related death at RM10 million. The weights c_ᴋ_, c_ƈ_, c_τ_, and c_Δ_ were set to 100, 2,400,000,000, 15,000, and 150, respectively, corresponding to self-protection costs of RM100 per day, Movement Control Order (MCO) 1.0 costs of RM2.4 billion per day, increased screening and contact tracing followed by quarantine costs of RM15,000 per day, and case detection followed by isolation costs of RM150 per day. The maximum control rates for increased screening and contact tracing followed by quarantine were set to 0.9, case detection followed by isolation was set to 0.3, movement control to 0.7, and self-protection to 0.25. The optimal control profile is shown in Figure 8. Figure 9 shows the forecasted trend of active cases (I_D_ and I_X_) and exposed cases (E_Q_ and E_X_), while Figure 10 shows the forecasted cumulative active cases (I_D__cum and I_X__cum) and deaths (D), if the optimal control strategy is implemented.

**Figure 7.**
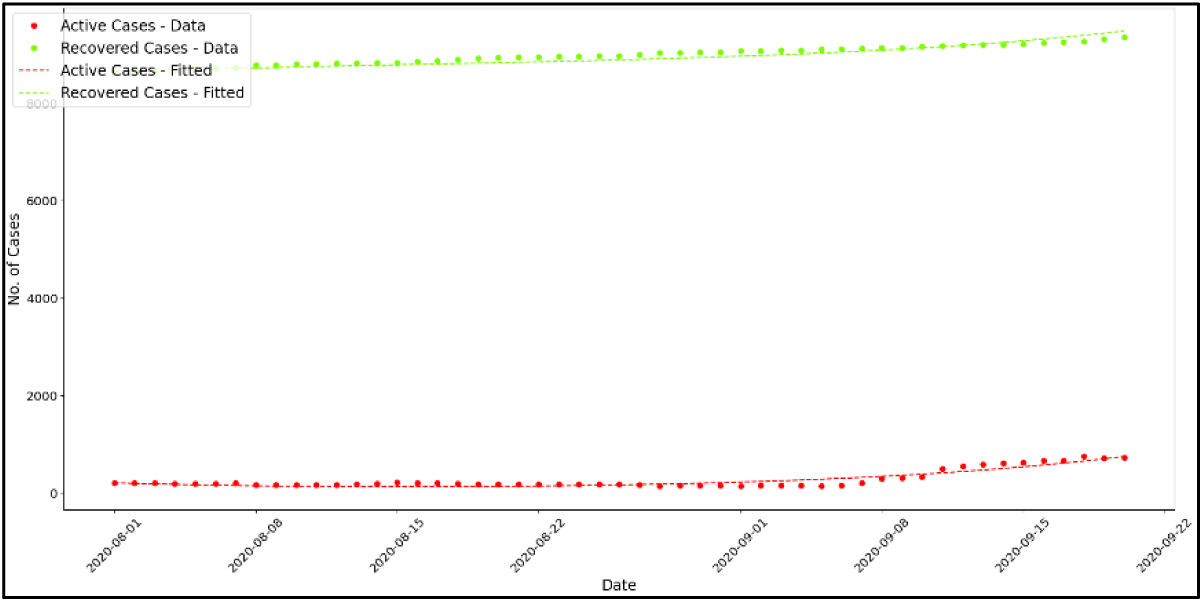
Curve Fitting Graph With The Introduction of Mandatory Mask Wearing

**Figure 8.**
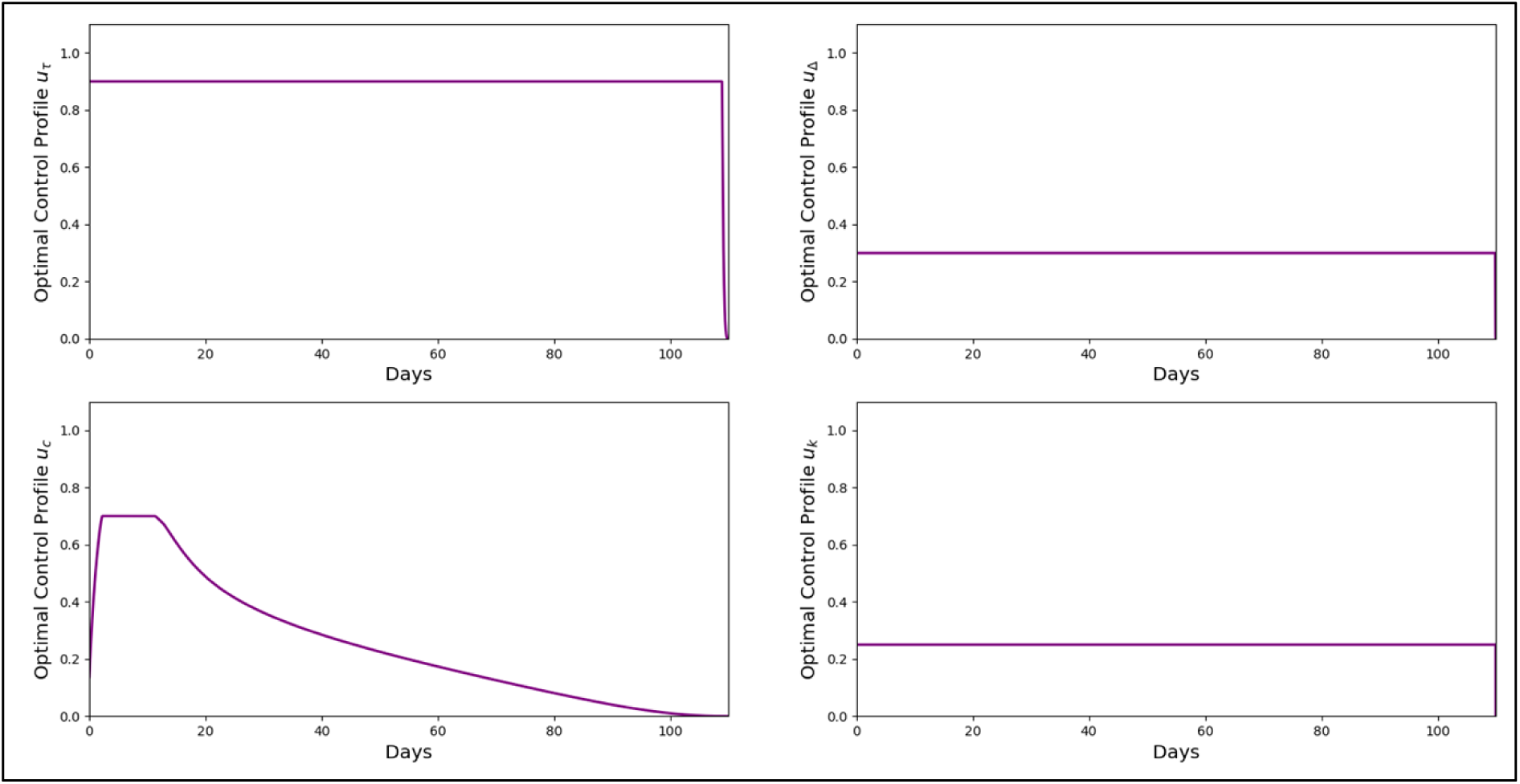
Optimal Control Profile for Simulation 1

**Figure 9.**
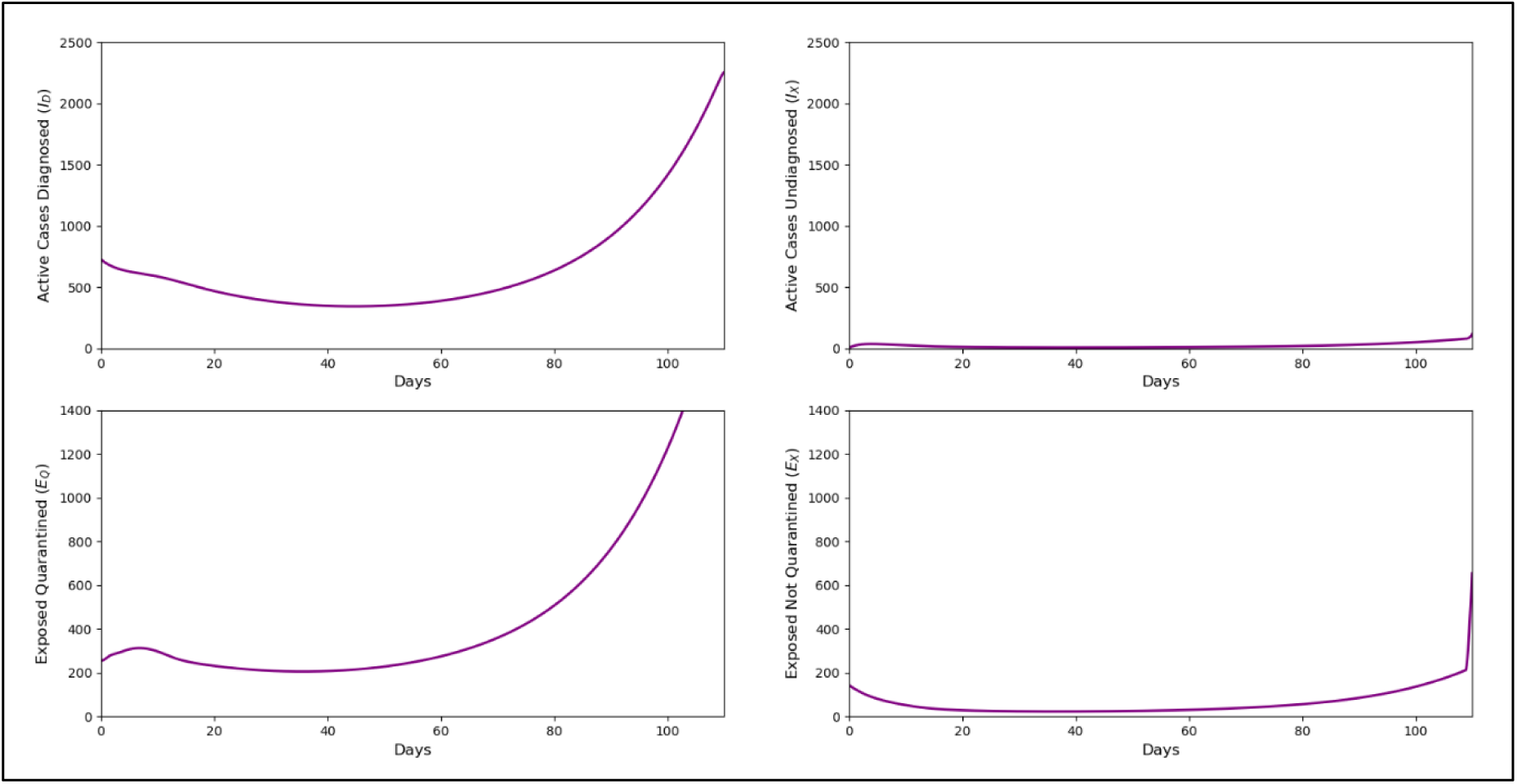
Forecasted trend of active cases (I_D_ and I_X_) and exposed cases (E_Q_ and E_X_) for Simulation 1

**Figure 10.**
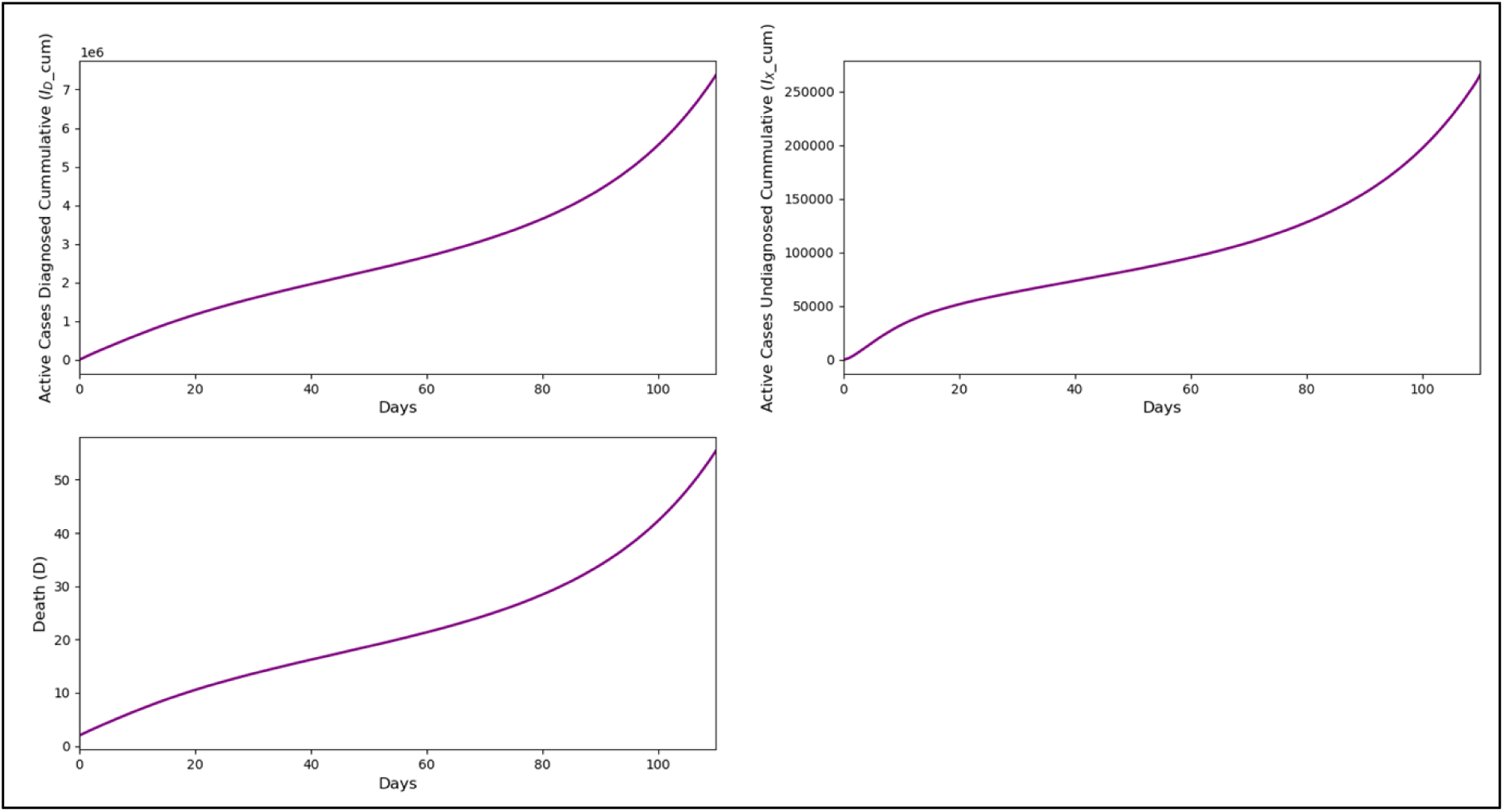
Forecasted trend of cumulative active cases (I_D__cum and I_X__cum) and deaths due to COVID-19 (D) for Simulation 1

In this simulation, with costs set to reflect real-world values, all four control strategies were activated at their maximum levels. However, the optimal control for movement control needed to decrease after approximately 10 days. This suggests that strict movement controls should only be maintained temporarily to avoid prolonged economic and social disruptions. Meanwhile, the other three controls remained at maximum level throughout, highlighting their importance in the long-term control of the pandemic. The trends for active cases (I_D_ and I_X_), exposed cases (E_Q_ and E_X_), and COVID-19-related deaths (D) all showed an increasing trend at the end of the 110-day forecast period. The number of diagnosed active cases (I_D_) was 2,255 on day 110, the number of undiagnosed active cases (I_X_) was 118, exposed cases quarantined (E_Q_) approached 1,300, and exposed cases not quarantined (E_X_) were about 250. The total number of deaths due to COVID-19 was found to be 55 on day 110.

#### Simulation 2: All Four Controls (u_ᴋ_, u_ƈ_, uτ, u_Δ_), with Enhanced Control Rate for uτ, and Costs Approximating Real Costs

By setting the weights for each infection, death, and control to the same values as in Simulation 1, but the maximum control rate for screening and contact tracing followed by quarantine increased to 0.95, while the maximum control rates for the other controls remained the same, the optimal control profile obtained is shown in Figure 11. Figure 12 and Figure 13 show the forecasted trend of active cases, exposed cases, and deaths, in comparison to Simulation 1 where the maximum control rate for screening and contact tracing followed by quarantine was 0.9.

**Figure 11.**
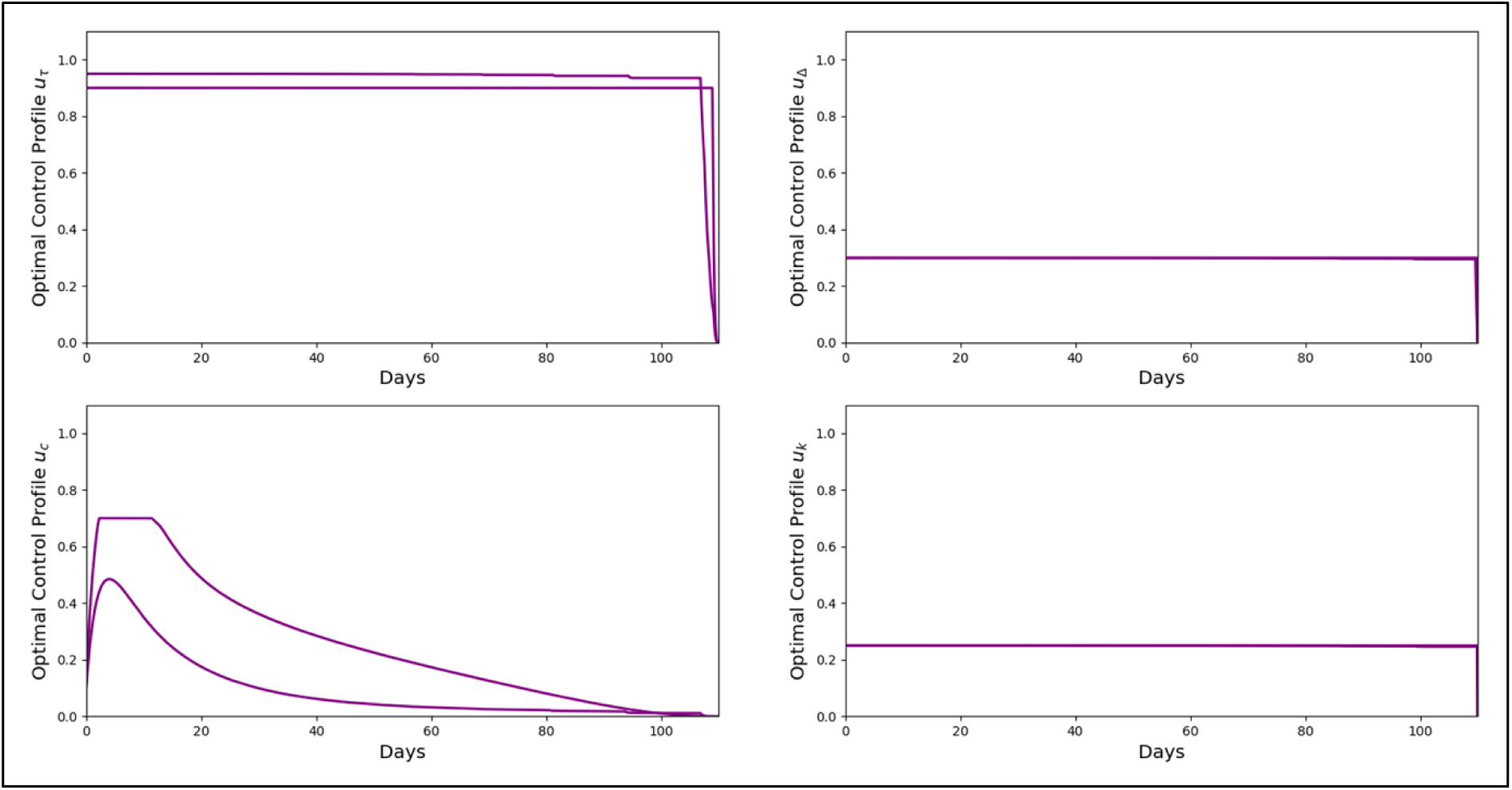
Optimal Control Profile for Simulation 2

**Figure 12.**
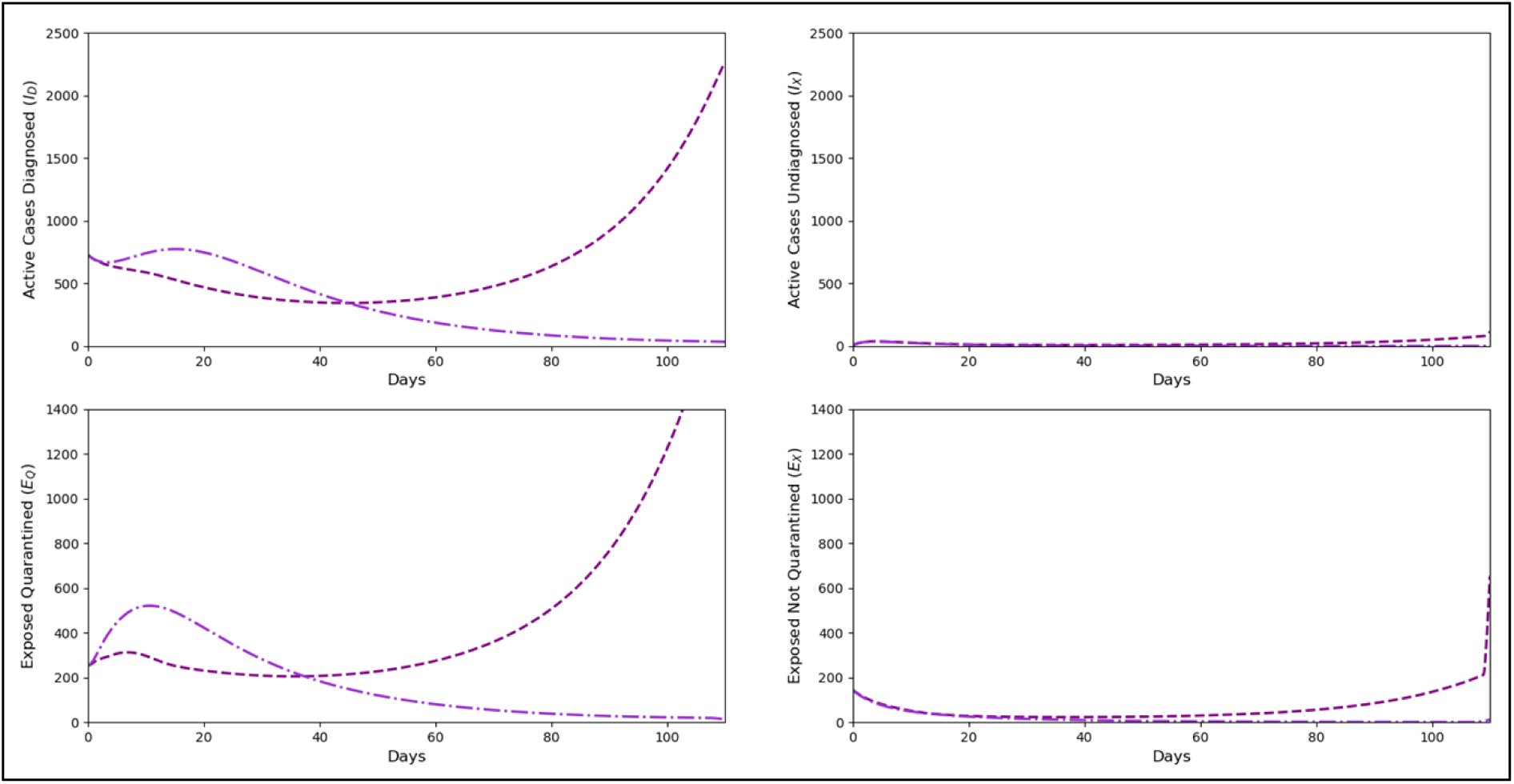
Forecasted trend of active cases (I_D_ and I_X_) and exposed cases (E_Q_ and E_X_) for Simulation 2

**Figure 13.**
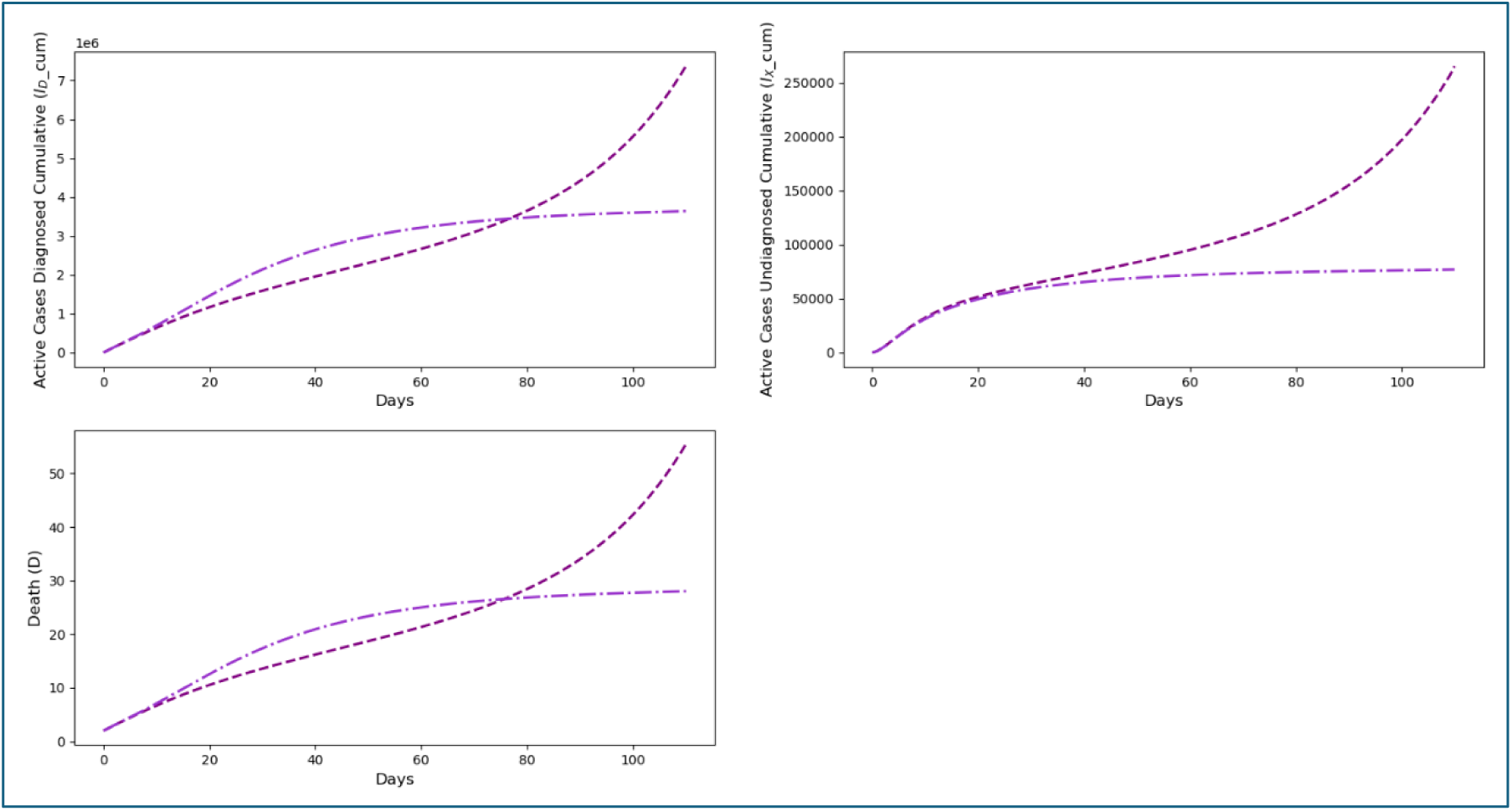
Forecasted trend of cumulative active cases (I_D__cum and I_X__cum) and deaths due to COVID-19 (D) for Simulation 2

When the control rate for screening and contact tracing (u_τ_) was increased to 0.95, the optimal control profile for movement control resulted in a shorter duration at maximum while the other three controls remained at maximum until the end of the forecast period. This simulation demonstrates that even with a shorter duration of maximum movement control, increasing the efficiency of screening and contact tracing can significantly impact the disease trajectory. The number of undiagnosed cases was low, not exceeding 37 cases per day. By the end of the forecast period, the cumulative diagnosed active cases (I_D__cum) were 3,640,480, almost half the reduction compared to Simulation 1. The number of deaths was found to have plateaued at no more than 30 cases on day 110 compared to 55 deaths at the end of the forecast period in Simulation 1.

#### Simulation 3: Three Controls (u_ᴋ_, u_ƈ_, u_Δ_), Without Increased Screening and Contact Tracing Followed by Quarantine (u_τ_), with Costs Approximating Real Costs

With the same weight values as in Simulations 1 and 2, the control profile for the intervention without screening and contact tracing followed by quarantine (u_τ_), is shown in Figure 14. Figure 15 and Figure 16 show the forecasted trend of active cases, exposed cases, cumulative active cases, and deaths if this control strategy is implemented.

**Figure 14.**
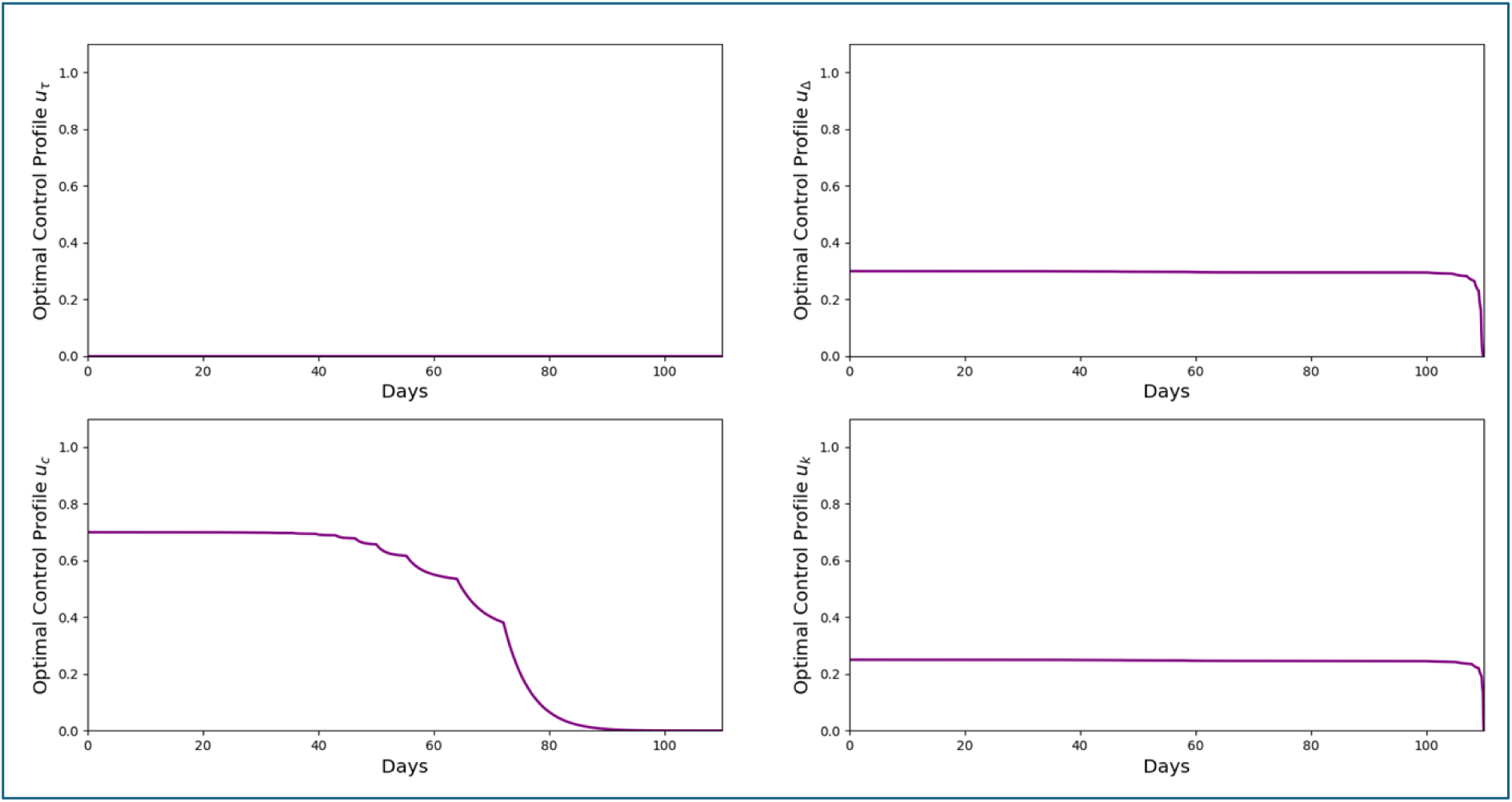
Optimal Control Profile for Simulation 3

**Figure 15.**
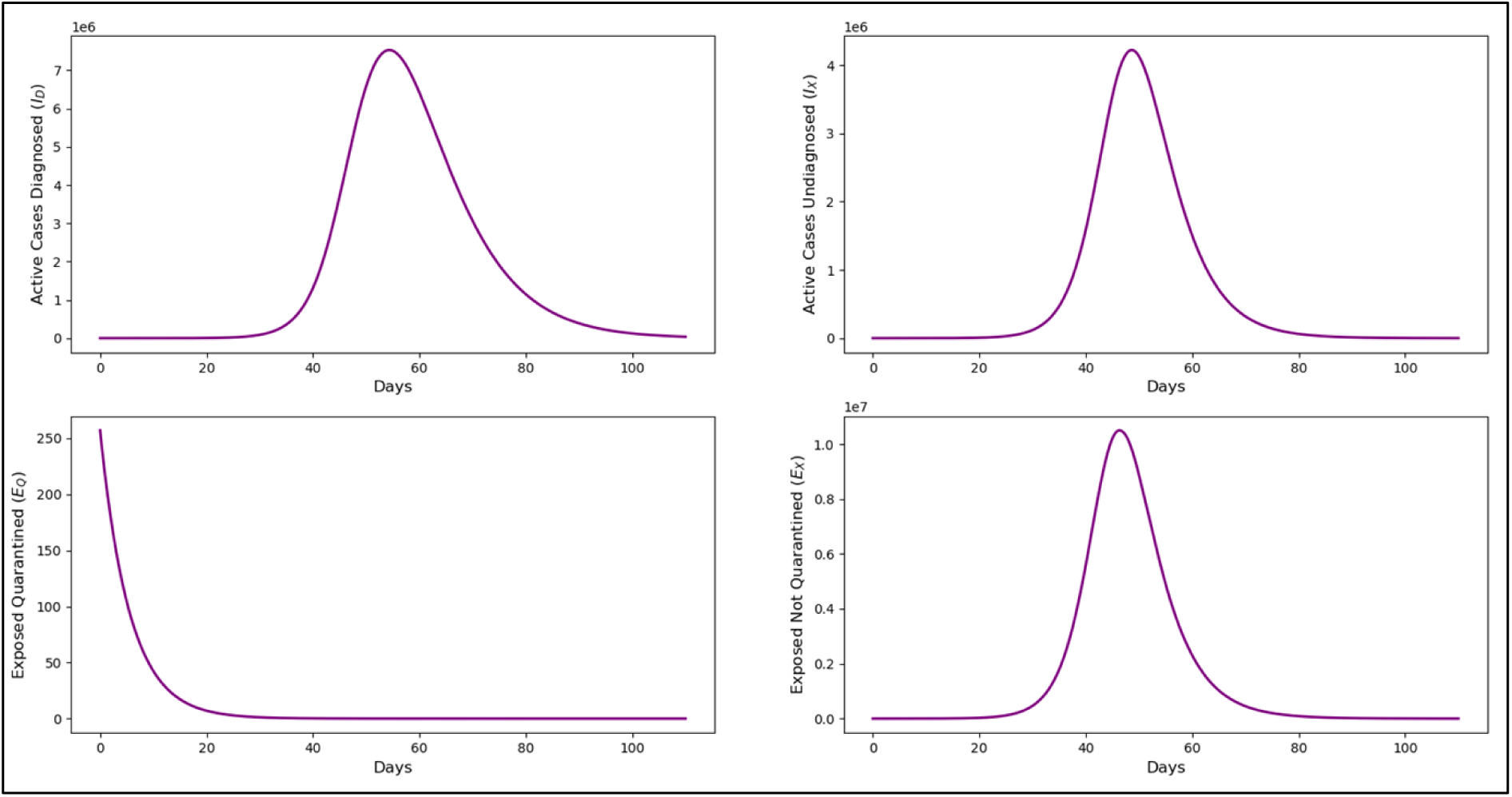
Forecasted trend of active cases (I_D_ and I_X_) and exposed cases (E_Q_ and E_X_) for Simulation 3

**Figure 16.**
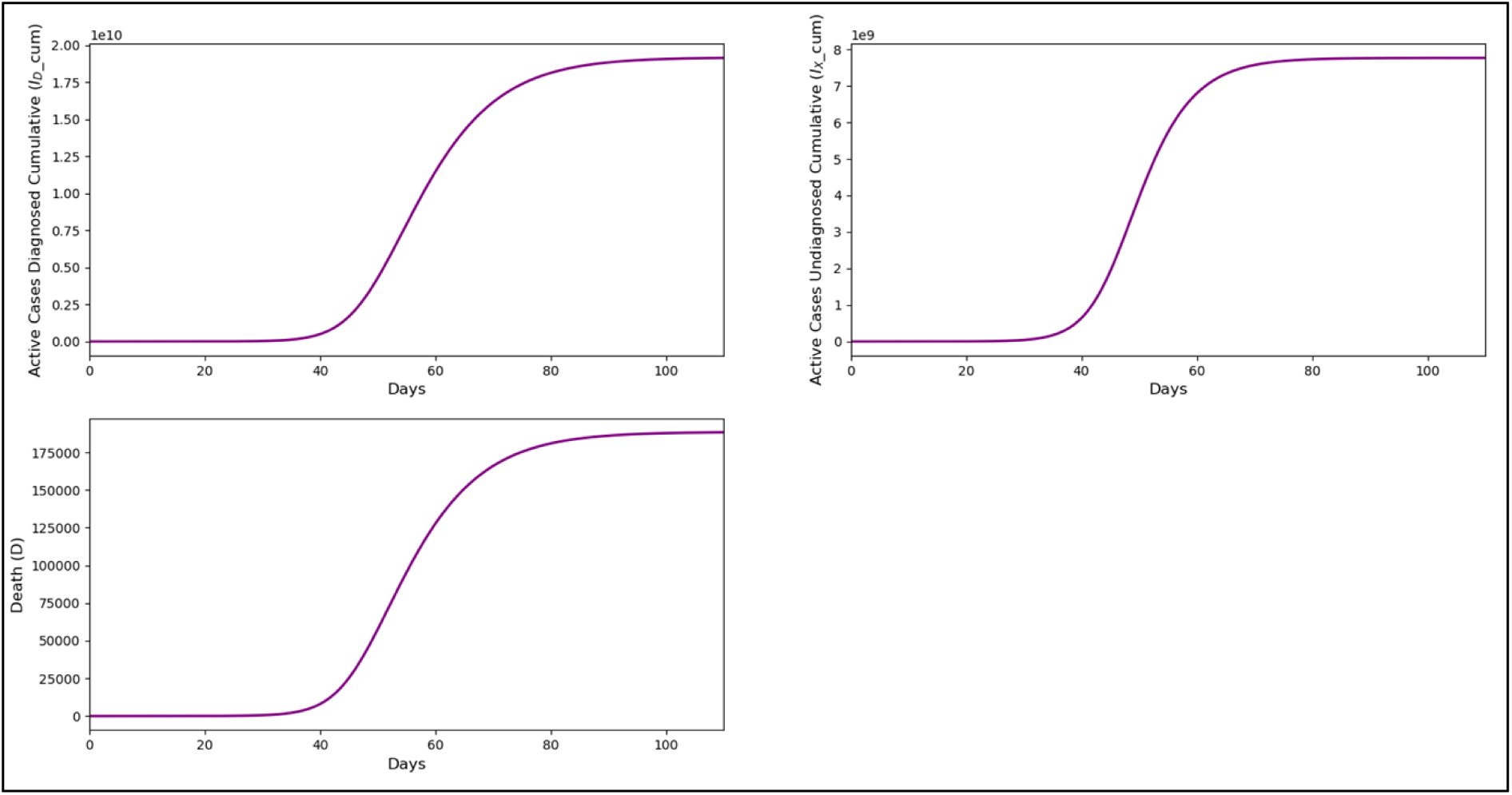
Forecasted trend of cumulative active cases (I_D__cum and I_X__cum) and deaths due to COVID-19 (D) for Simulation 3

Without screening and contact tracing followed by quarantine (u_τ_), the optimal control profile for self-protection (u_ᴋ_) and case detection followed by isolation (u_Δ_) remained at maximum until the end of the forecast period, while movement control (u_ƈ_) began to decrease after approximately 40 days at the maximum, approaching zero by day 90. This was found to cause a sharp increase in active cases with the number of diagnosed active cases reached up to 7,524,906 cases per day, while the undiagnosed active cases reached 4,221,351 cases per day. The cumulative diagnosed active cases (I_D__cum) exceeded 19 billion, and the cumulative undiagnosed active cases (I_X__cum) exceeded 7 billion at the end of the forecast period. The total number of deaths due to COVID-19 by the end of the forecast period was 188,295. The trend of infected individuals reached its peak around day 50 and then declined, possibly due to the decreasing number of susceptible individuals (S) approaching zero at that time. This simulation underscores the critical role of contact tracing in controlling outbreaks. Without effective screening, even other interventions like self-protection and movement control were insufficient to prevent a large-scale surge in cases.

#### Simulation 4: Three Controls (u_ᴋ_, u_τ_, u_Δ_), Without Movement Control (u_ƈ_), with Costs Approximating Real Costs

With the same weight values as in Simulations 1 and 3, the optimal control profile for the intervention excluding movement control (u_ƈ_) is shown in Figure 17. Figure 18 and Figure 19 show the forecasted trend of active cases, exposed cases, cumulative active cases, and deaths, if this control strategy is implemented.

**Figure 17.**
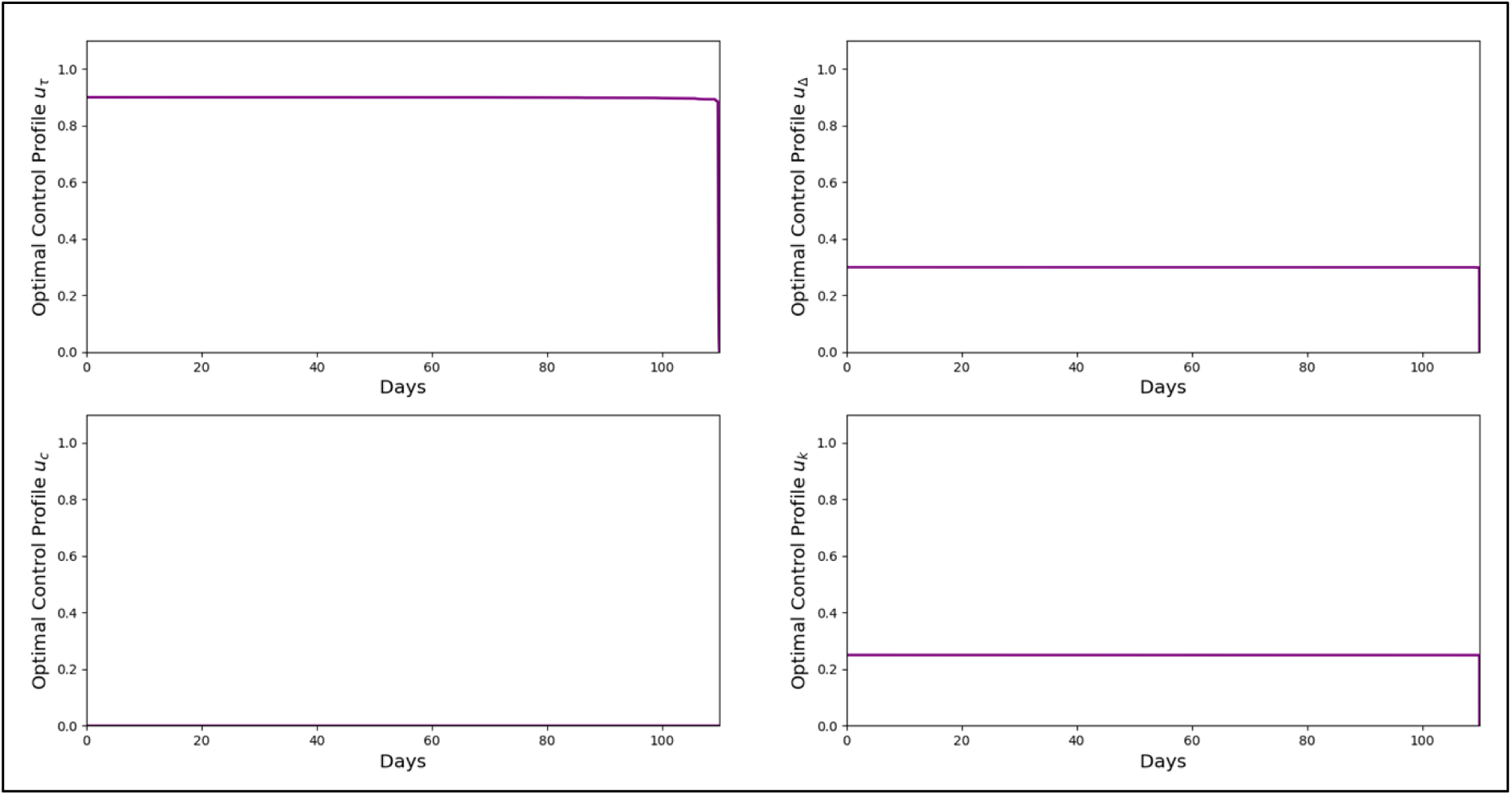
Optimal Control Profile for Simulation 4

**Figure 18.**
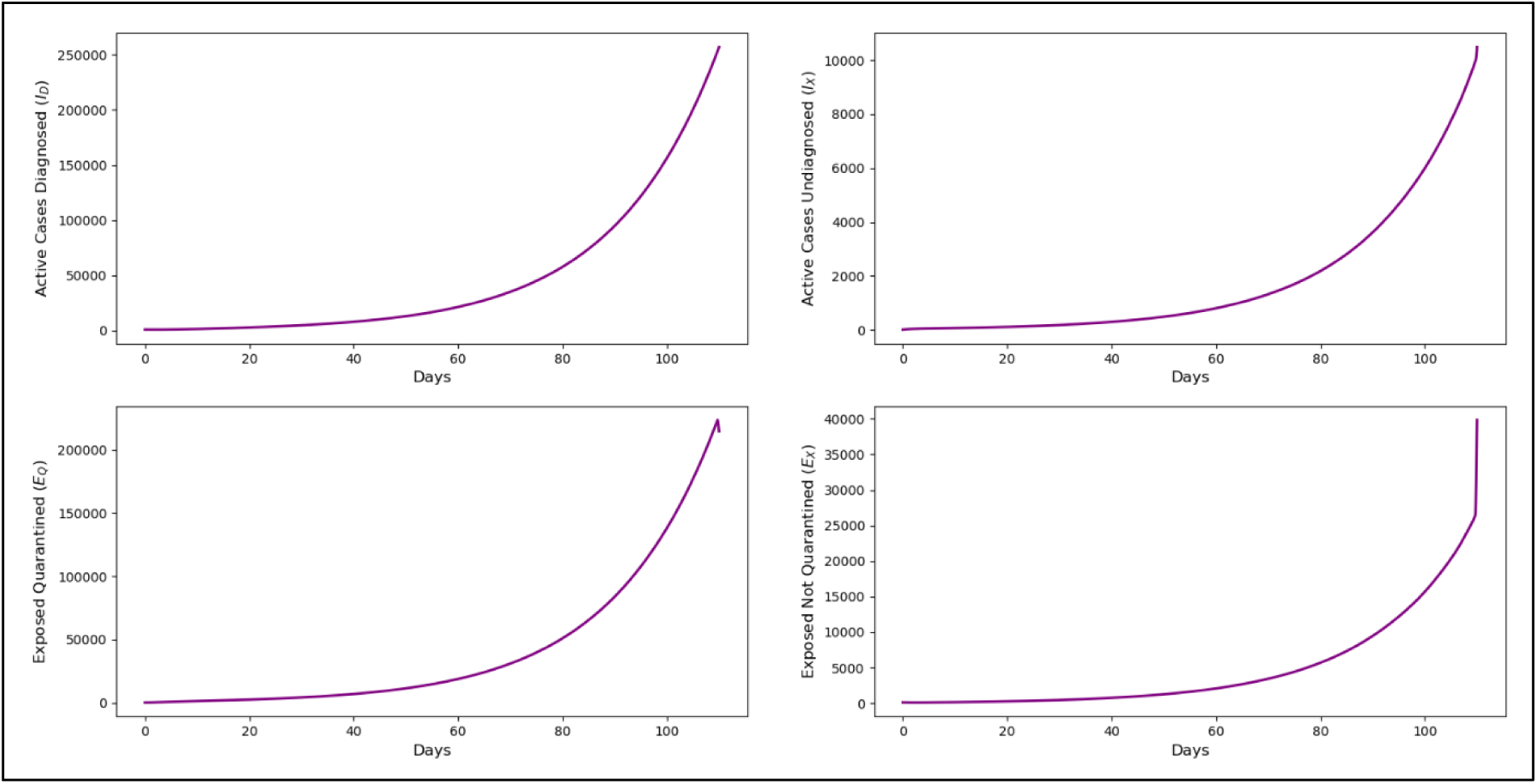
Forecasted trend of active cases (I_D_ and I_X_) and exposed cases (E_Q_ and E_X_) for Simulation 4

**Figure 19.**
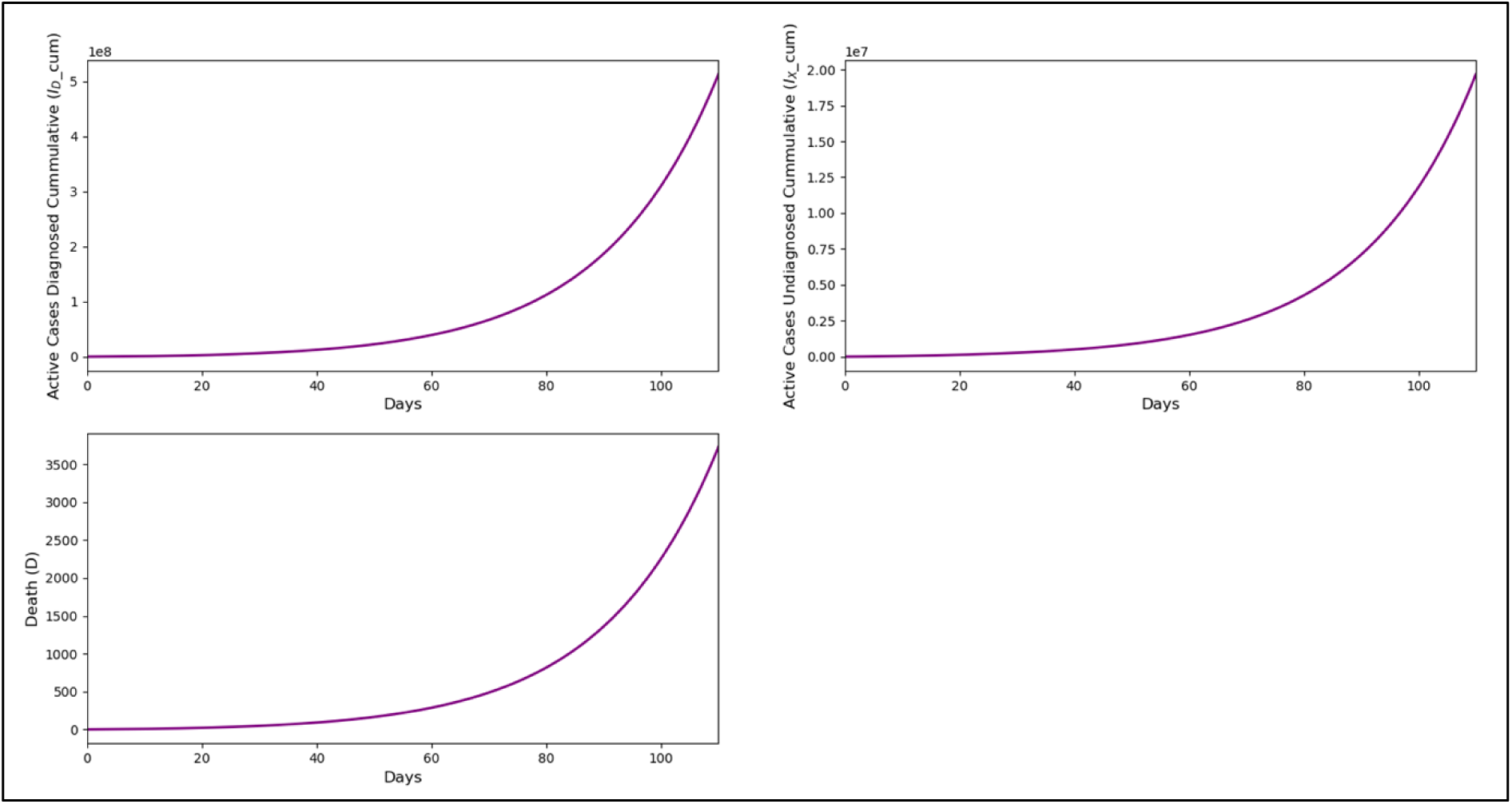
Forecasted trend of cumulative active cases (ID_cum and IX_cum) and deaths due to COVID-19 (D) for Simulation 4

Without movement control (u_ƈ_), the optimal control profile for the other three controls remains at maximum levels until the end of the forecast period. With this strategy, the trends for infected, exposed, and deaths were found to still be on the rise at the end of the forecast period. On day 110, the number of diagnosed active cases (I_D_) peaked at 256,923 cases per day, and 512,666,535 cumulatively (I_D__cum). The number of undiagnosed active cases (I_X_) reached a maximum of more than 10 thousand cases per day, and 19,665,355 cumulatively (I_X__cum). The total number of deaths due to COVID-19 by the end of the forecast period was approximately 4,000.

#### Simulation 5: Strategy with All Four Control Types (u_ᴋ_, u_ƈ_, u_τ_, u_Δ_), with Decrease in c_ƈ_ (Cost of u_ƈ_)

By setting the weights for each infection, death, and control except for movement control to the same values as in Simulations 1 through 4, and keeping the maximum control levels the same, Figure 20 shows the optimal control profile with varying weights for movement control (c_ƈ_). Four weight values were used, starting with 2,400,000,000, as the original value in Simulation 1 corresponding to the real cost of MCO 1.0 which is RM2.4 billion per day, followed by 1,000,000,000, 100,000,000, and finally 10,000,000. Figure 21 and Figure 22 show the forecasted trend of active cases, exposed cases, cumulative active cases, and deaths, if this control strategy is implemented.

**Figure 20.**
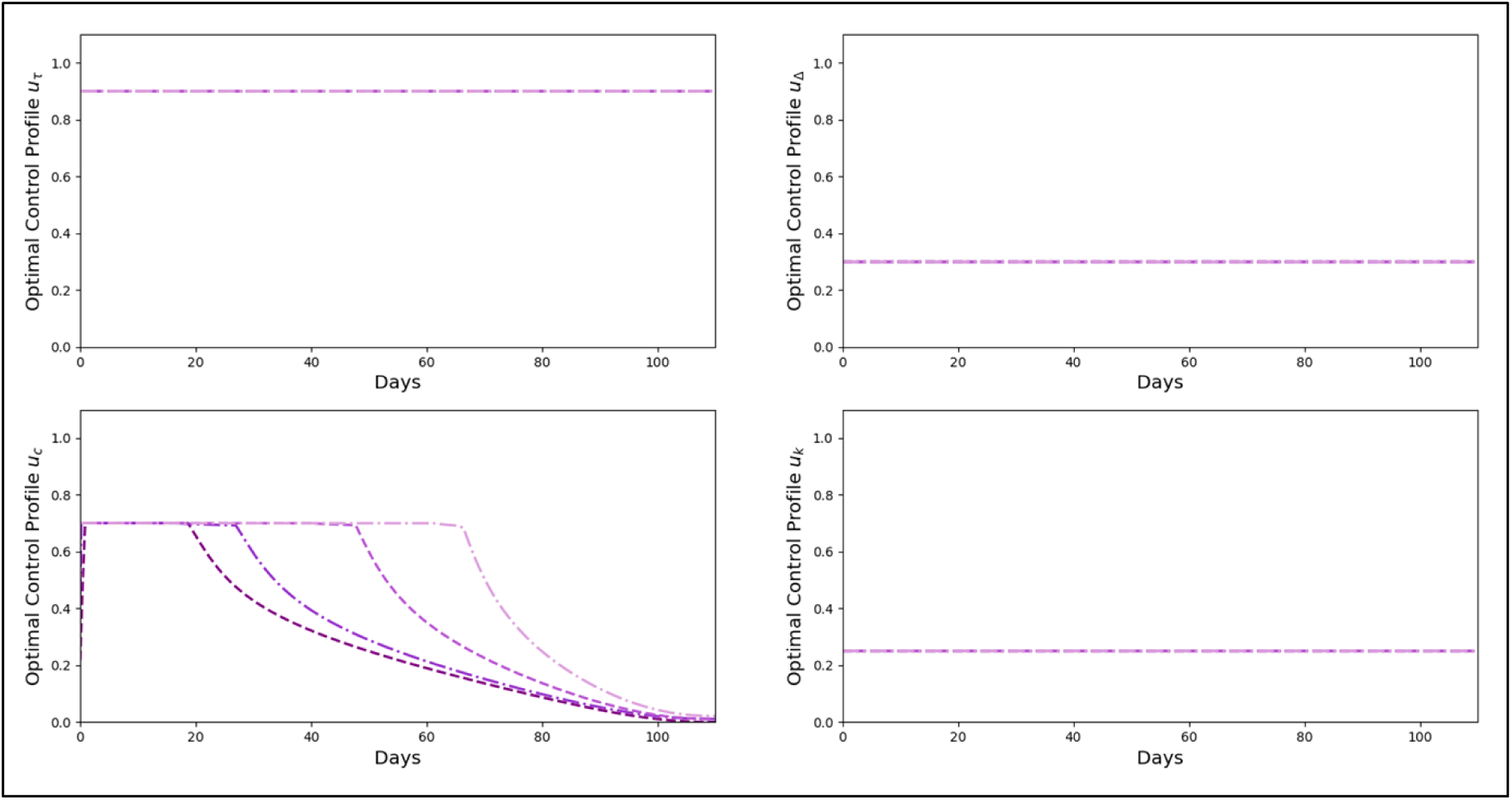
Optimal Control Profile for Simulation 5

**Figure 21.**
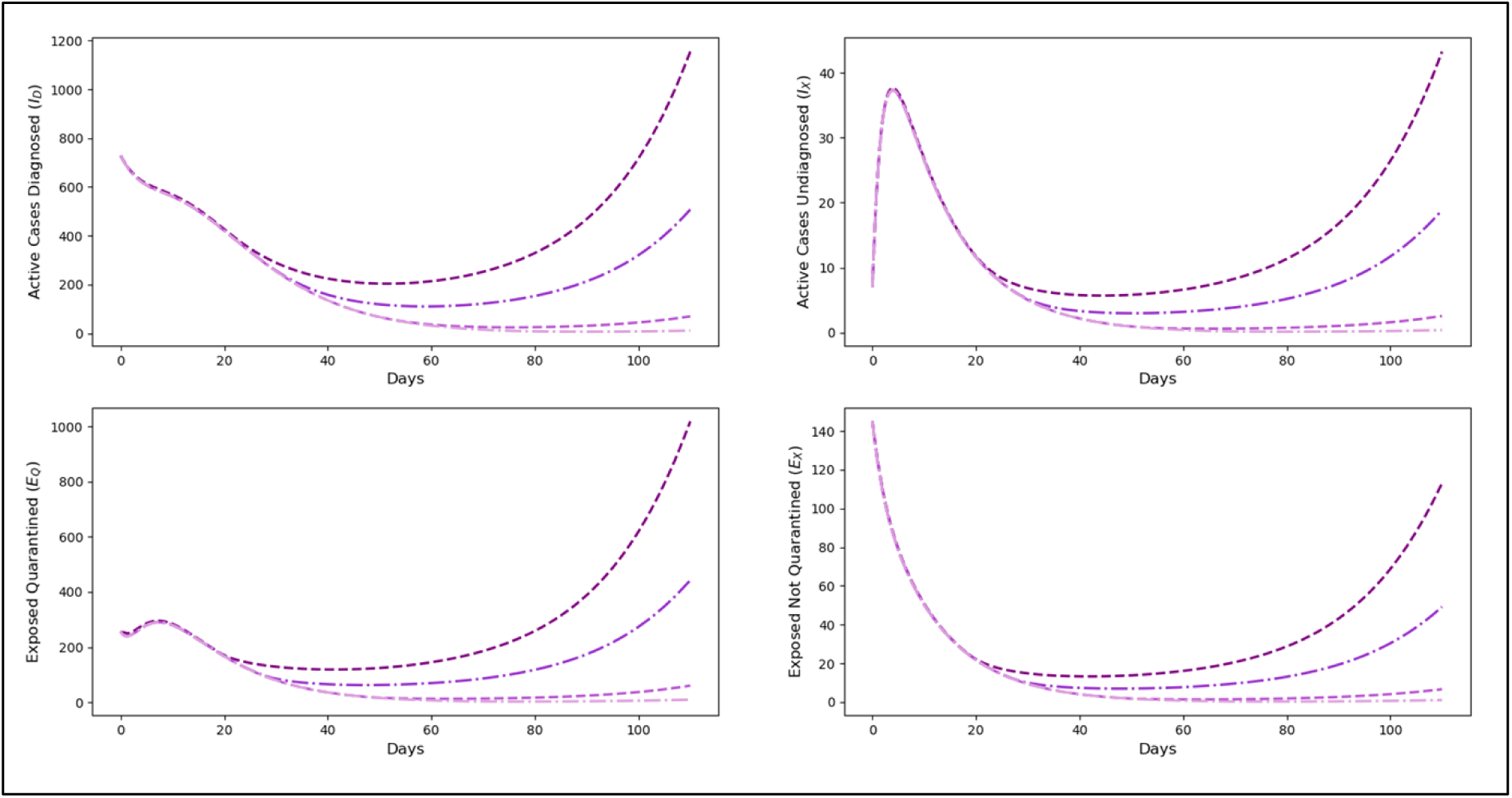
Forecasted trend of active cases (ID and IX) and exposed cases (EQ and EX) for Simulation 5

**Figure 22.**
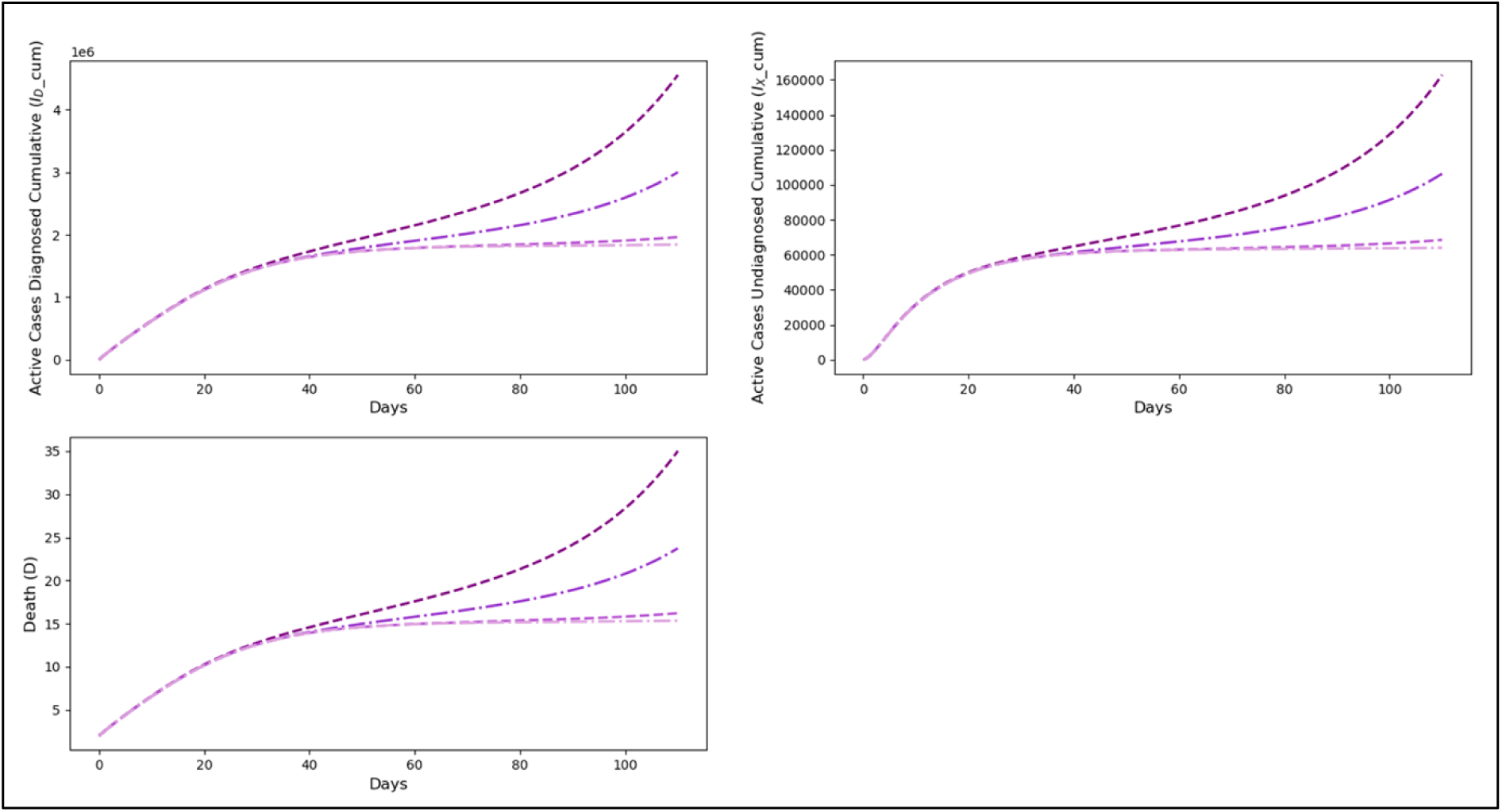
Forecasted trend of cumulative active cases (ID_cum and IX_cum) and deaths due to COVID-19 (D) for Simulation 5

By changing the weight values for movement control (c_ƈ_), the resulting optimal movement control profile was altered, but the optimal profiles for the other three controls remained at maximum levels until the end of the forecast period. The movement control profile was found to be at the maximum level, but the duration at maximum was longer with lower weight values. With a lower weight for movement control (c_ƈ_), the duration of maximum movement control was extended, thus achieving faster control over the trends in infections and deaths.

#### Simulation 6: Strategy with All Four Control Types (u_ᴋ_, u_ƈ_, u_τ_, u_Δ_), with Varying Maximum Movement Control Rate (u_ƈ_)

Simulation 6 was conducted with the weights for each infection and death set to the same values as in Simulations 1 through 4. However, unlike Simulation 5, the reduction in the weight for movement control was aligned with the reduction in the maximum movement control rate. Figure 23 shows the optimal control profile for varying maximum movement control rates, with the weight for movement control (c_ƈ_) set to 300,000,000, corresponding to the economic loss during MCO 2.0, which was RM300 million per day. Three maximum control levels were set, which are 0.7, as the level obtained during MCO 1.0, followed by 0.6, and 0.5, which are close to the control level obtained during MCO 2.0. Figure 24 and Figure 25 show the forecasted trend of active cases, exposed cases, cumulative active cases, and deaths, if this control strategy is implemented.

**Figure 23.**
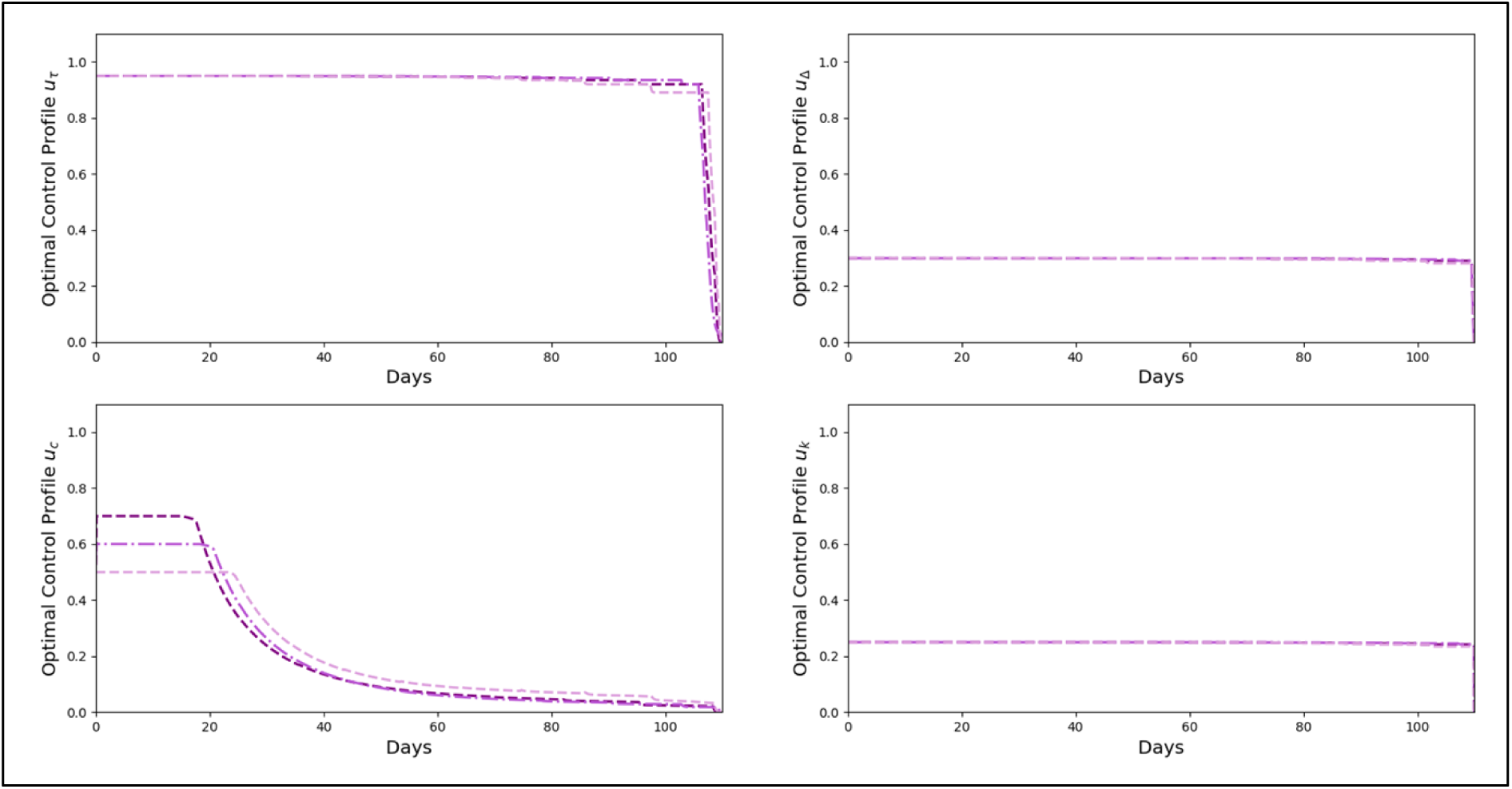
Optimal Control Profile for Simulation 6

**Figure 24.**
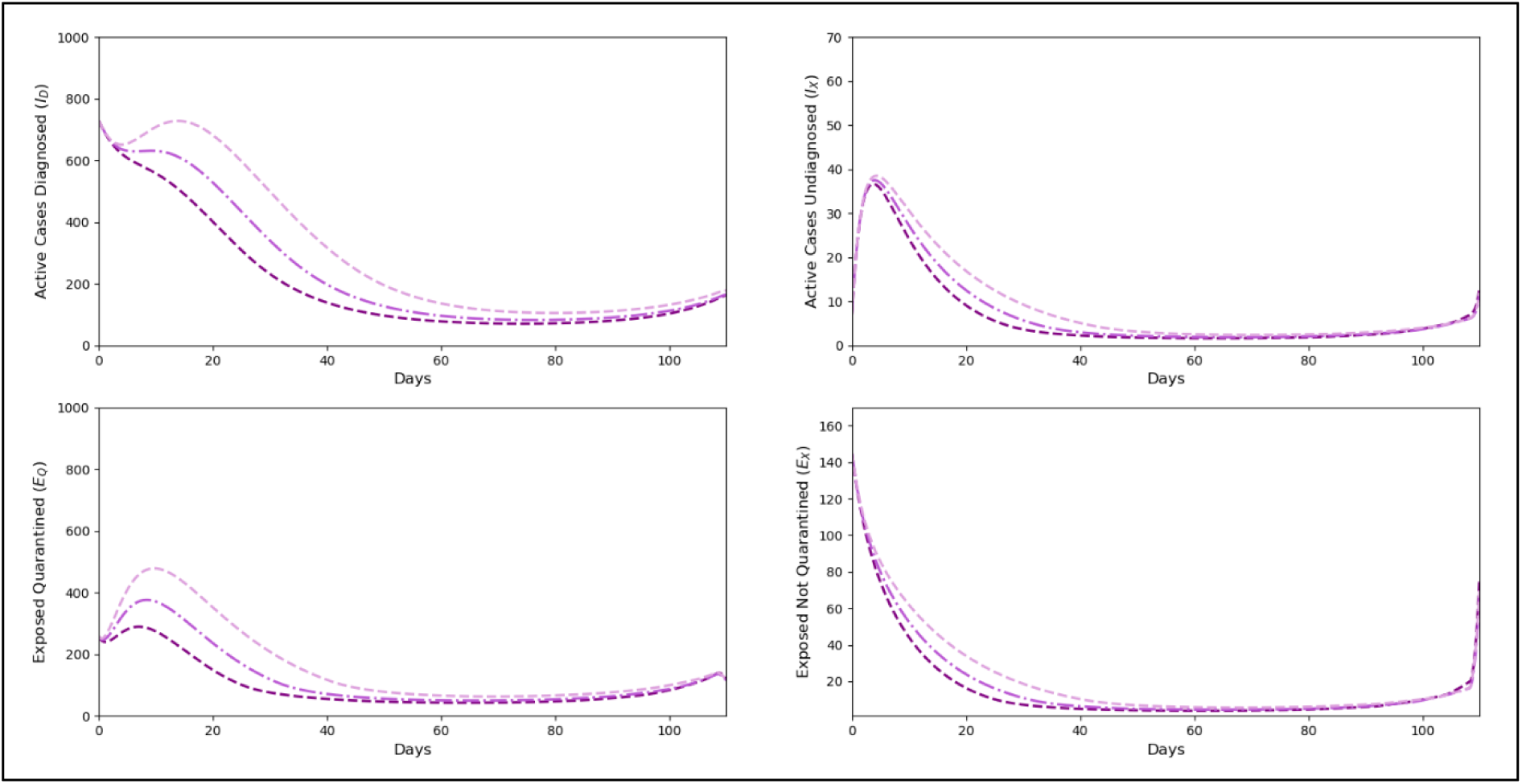
Forecasted trend of active cases (I_D_ and I_X_) and exposed cases (E_Q_ and E_X_) for Simulation 6

**Figure 25.**
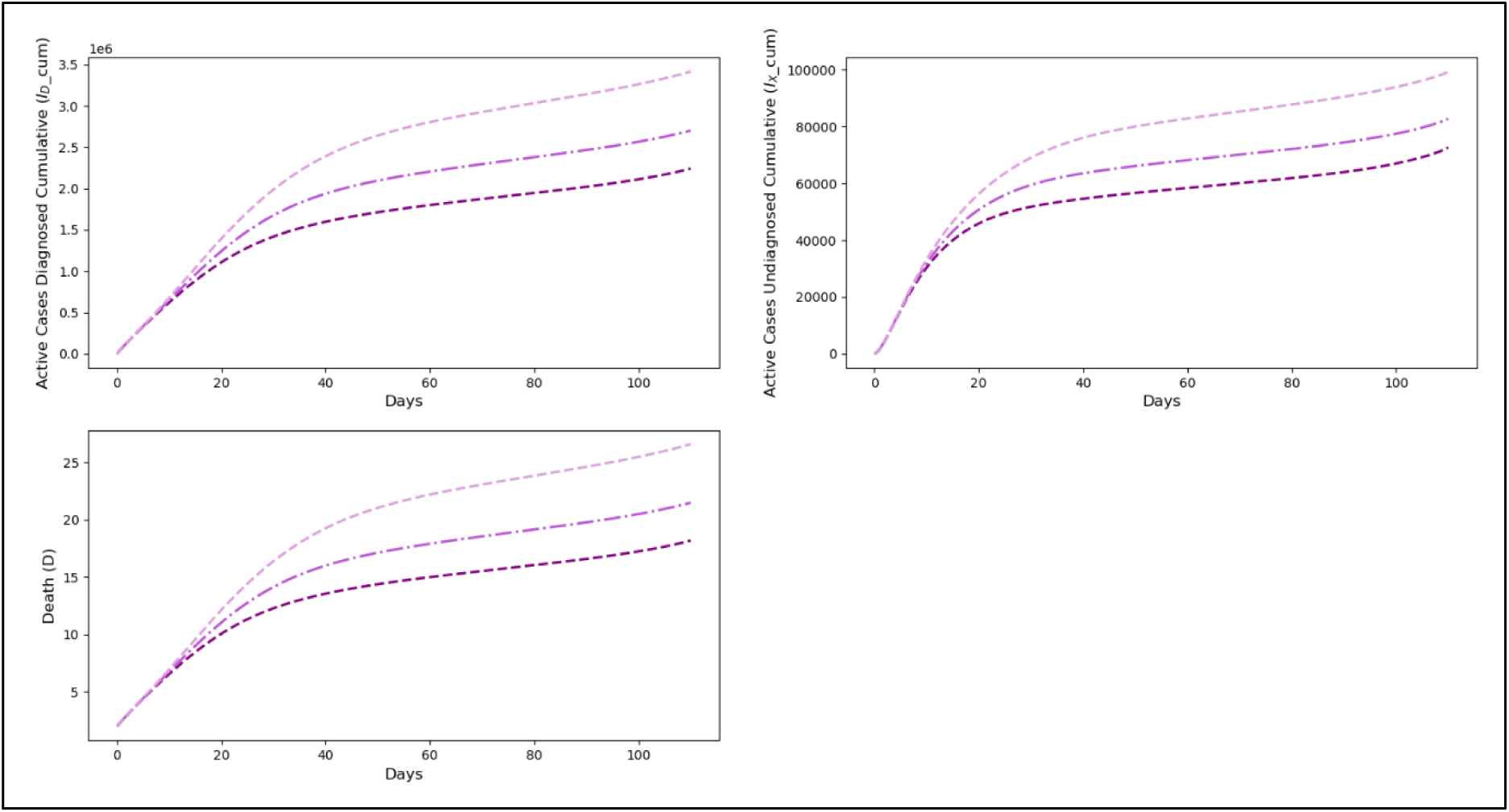
Forecasted trend of cumulative active cases (I_D__cum and I_X__cum) and deaths due to COVID-19 (D) for Simulation 6

Simulation 6 differs from Simulation 5 in that the reduction in the cost during MCO matches with reduction in the control level, which is realistic in a real-world setting. Through this optimal control modelling, by altering the maximum rate of movement control (u_ƈ_), the resulting optimal movement control profile was changed, but the optimal profiles for the other three controls remained at maximum levels until the end of the forecast period. The generated movement control profile was found to be at each maximum level, but the duration at the maximum level was longer with lower control rates. Trends for infected cases, exposed, and deaths were more quickly controlled with higher control rates, even if the duration of the control was shorter. The cumulative active cases and deaths were also lower with stricter controls. However, the differences in the numbers were seen not overly significant. The cumulative diagnosed active cases (I_D__cum) at the end of the forecast period were 2,241,486 at a control rate of 0.7, 2,702,292 at 0.6, and 3,415,396 at 0.5. The cumulative undiagnosed active cases (I_X__cum) at the end of the forecast period were 72,686 at a control rate of 0.7, 82,773 at 0.6, and 99,371 at 0.5. The total number of deaths was 18 at a control rate of 0.7, 21 at 0.6, and 27 at 0.5.

#### Simulation 7: Strategy with All Four Control Types (u_ᴋ_, u_ƈ_, u_τ_, u_Δ_), with Varying Maximum Movement Control Rate (u_ƈ_)

In Simulations 1 through 6 above, the optimal control modelling analysis was conducted by incorporating diagnosed active cases, undiagnosed active cases, and COVID-19-related deaths into the objective function. In the subsequent Simulation 7, however, optimal control modelling was conducted by considering only active cases (both diagnosed and undiagnosed) in the objective function, without accounting for deaths.

The resulting control profiles from this modelling are shown in Figure 4.77, while the projected trends for cumulative active cases and COVID-19 deaths are shown in Figure 4.78, and compared with a similar scenario—Simulation 1 in Section 4.9.1a.

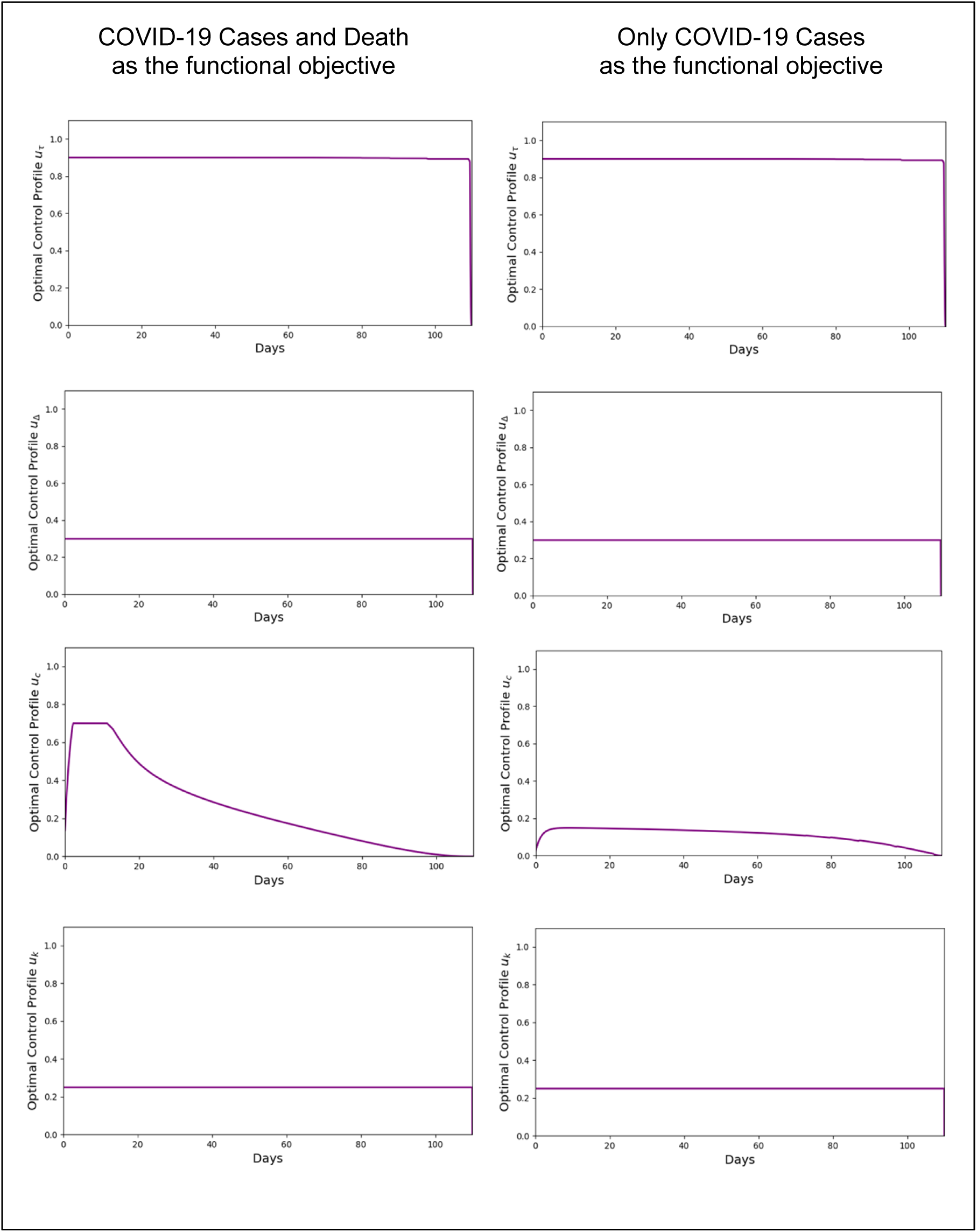

When only active cases were considered, with a daily cost of RM4,000 represented by its weight in the objective function, and deaths were excluded (each previously assigned a cost of RM10 million), the resulting control profiles were similar except for the movement restriction control. The movement control rate obtained in this simulation did not reach the maximum level of 0.7, but instead remained at a lower level of approximately 0.2 and persisted until near the end of the projection period. The resulting trends in active cases and deaths were significantly higher when the number of deaths was excluded from the objective function, compared to when it was included.

In this study, optimal control theory is employed to identify the most effective control strategies for reducing the spread of COVID-19 within a population, while minimizing the impacts on the social, economic, and public health sectors. Optimal control theory involves the dynamic adjustment of interventions such as movement restrictions, physical distancing, vaccination, and health screening within a mathematical model, in order to determine control strategies that optimize outcomes across various dimensions.

In this context, the use of the SEIR model with optimal control allows for simulations to assess the long-term effects of the control measures implemented, with the objective of minimizing the negative impacts on the population. For example, measures such as border closures and movement restrictions may reduce infection rates, but they also have significant repercussions on the economy and social stability. Through optimal control theory, this model can determine a balanced strategy, where the control measures effectively curb the spread of the disease, while simultaneously minimizing disruptions to economic and social activities, and maintaining the capacity of the health system to avoid being overwhelmed.

In the realm of public health, optimal control theory also aids in the more efficient allocation of health resources. For instance, this model can evaluate how vaccination strategies or gradual economic reopening can be managed to reduce the burden on the healthcare system (such as lowering ICU admissions) without triggering a sharp rise in virus transmission. By determining the optimal strategy, optimal control theory supports adaptive and flexible policy-making, providing recommendations based on relevant data and simulations.

## 4. Conclusion

This study primarily focuses on the development of a mathematical epidemiological model of COVID-19 in Malaysia, utilizing the SEIR (Susceptible, Exposed, Infectious, Recovered) approach to provide a comprehensive understanding of disease transmission dynamics and the efficacy of control interventions on epidemiological trends. A structured mathematical methodology was employed, incorporating relevant variables and accurate parameters to explore the characteristics of COVID-19, including its transmission factors and potential impacts.

Through the analysis of the SEIR model developed in this study, a clearer depiction of the nature of COVID-19 has been achieved, serving as a critical indicator for understanding the infectious potential of the disease. Additionally, this model effectively illustrates the impact of health interventions on disease trends. The model’s ability to predict fluctuating epidemiological patterns, influenced by changes in public health policies and strategies, highlights its importance in assessing disease control strategies.

Through the application of optimal control theory within the SEIR model, this study offers more targeted and effective solutions for controlling the spread of COVID-19. It enables the identification of control measures that can reduce negative impacts across all dimensions, including social, economic, and health aspects, while maintaining a balance between the need to control the outbreak and the necessity of ensuring the continuity of economic and social functions. This makes the model a highly valuable tool in the formulation of more effective and sustainable disease control policies and strategies.

The insights gained from this modelling offer significant implications for public health policy, particularly in shaping long-term strategies to combat prolonged pandemics. The model’s capacity to accurately reflect the epidemiological trends of COVID-19 in Malaysia across various conditions, time phases, and scenarios underscores its validity and reliability. This study has demonstrated that the SEIR mathematical model can serve as a powerful tool for future decision-making related to the control of infectious diseases. The findings advocate for the continued use of mathematical models in public health planning, underscoring their role in guiding interventions and forecasting disease behaviour in response to policy changes.

## Acknowledgement

None

**The authors declare no conflicts of interest**

## Data Availability Statement

All data are in the manuscript and/or supporting information files

